# Sewer monitoring for antimicrobial resistance genes and organisms at healthcare facilities

**DOI:** 10.1101/2025.03.16.25324079

**Authors:** Rachel Poretsky, Dolores Sanchez Gonzalez, Adam Horton, Michael Schoeny, Chi-Yu Lin, Modou Lamin Jarju, Michael Secreto, Cecilia Chau, Ellen Gough, Erin Newcomer, Adit Chaudhary, Lisa Duffner, Nidhi Undevia, Angela Coulliette-Salmond, Amanda K. Lyons, Florence Whitehill, Mary K. Hayden, Stefan J. Green, Michael Y. Lin

## Abstract

Surveillance of wastewater from healthcare facilities has the potential to identify the emergence of multidrug-resistant organisms (MDROs) of public health importance. Specifically, wastewater surveillance can provide sentinel surveillance of novel MDROs (e.g., emergence of *Candida auris*) in healthcare facilities and could help direct targeted prevention efforts and monitor longitudinal effects. Several knowledge gaps need to be addressed before wastewater surveillance can be used routinely for MDRO surveillance, including determining optimal approaches to sampling, processing, and testing wastewater for MDROs. To this end, we evaluated multiple methods for wastewater collection (passive, composite, and grab), concentration (nanoparticles, filtration, and centrifugation), and PCR quantification (real-time quantitative PCR vs. digital PCR) for *C. auris* and 5 carbapenemase genes (*bla_KPC_, bla_NDM_, bla_VIM_, bla_IMP_*, and *bla_OXA-48-like_*) twice weekly for 6 months at a long-term acute care hospital in Chicago, IL. We also tested the effects of different transport and sample storage conditions on PCR quantification. All genes were detected in facility wastewater, with *bla_KPC_* being the most consistently abundant. Experiments were done in triplicate with gene copy, variance, and number of detections between triplicates used to determine method efficacy. We found that passive samples processed immediately using a combination of centrifugation followed by bead-beating and dPCR provided the most reliable results for detecting MDROs. We also present the trade-offs of different approaches and use culture and metagenomics to elucidate clinical relevance.

This study establishes a practical approach for wastewater surveillance as a potential tool for public health monitoring of MDRO burden in healthcare facilities.

## Background

Surveillance of wastewater from healthcare facilities has the potential to identify the emergence of multidrug-resistant organisms (MDROs) of public health importance [1]. Optimal approaches to sampling, processing, and testing wastewater need to be established before wastewater surveillance (WWS) can be used routinely for MDRO surveillance. Although genes of interest can be detected using similar molecular approaches in both wastewater samples and patient samples, there are multiple interrelated and unique challenges to WWS. First, wastewater is a complex medium, containing a mixture of organic matter, particles, microorganisms, and flow variability. Second, laboratory methods for WWS have not yet been standardized, leading to poor understanding of measurement uncertainties and the effects of wastewater composition on detection [2–4]. Finally, target abundance can hinder detection; previous metagenomics-based studies in municipal and hospital wastewater have shown that many antimicrobial resistance (AR) genes are present at low relative abundances [5–7].

Here, we present an evaluation of approaches to address these knowledge gaps, focusing on healthcare associated pathogens: *Candida auris* and carbapenemase-producing organisms (CPOs, represented by five target genes *bla_KPC_, bla_NDM_, bla_VIM_, bla_IMP_*, and *bla_OXA-48-like_*). We collected wastewater from a region of the United States where *C. auris* and CPOs are endemic [8–11]. This study employed a longitudinal design at a single long-term acute care hospital over a period of 6 months. We compared three sample collection methods (passive, composite, and grab), three sample processing methods (magnetic nanoparticle [NP] concentration, InnovaPrep Concentrating Pipette [ICP] filtration, and centrifugation followed by bead-beating), and two different detection methods (quantitative real-time PCR [qPCR] and digital PCR [dPCR]) with MDRO diagnostic assays that have been developed and validated for use in clinical samples; therefore, additional evaluation is needed to optimize performance in wastewater samples. We report the findings of a series of seminal experiments that establish an effective protocol for surveillance of *C. auris* and CPOs in healthcare facility wastewater, helping to define the use of WWS as an important tool for monitoring MDRO burden in healthcare facilities.

## Methods

### Sample collection

Wastewater samples were collected at an 86-bed long-term acute care hospital located in Chicago, Illinois (baseline data, Table S1). We collected samples twice weekly from June-December 2023 using three different collection methods: passive (Moore swabs consisting of sterile cotton gauze; n=50 collections yielding 149 samples), grab (n=50 collections yielding 288 samples), and composite (n=50 collections yielding 299 samples) (Fig. 1A, Supplemental Materials) from a manhole that represents entire facility (per a dye test, Supplemental Materials). Each sample was homogenized before being subsampled in triplicate.

**Figure 1.**
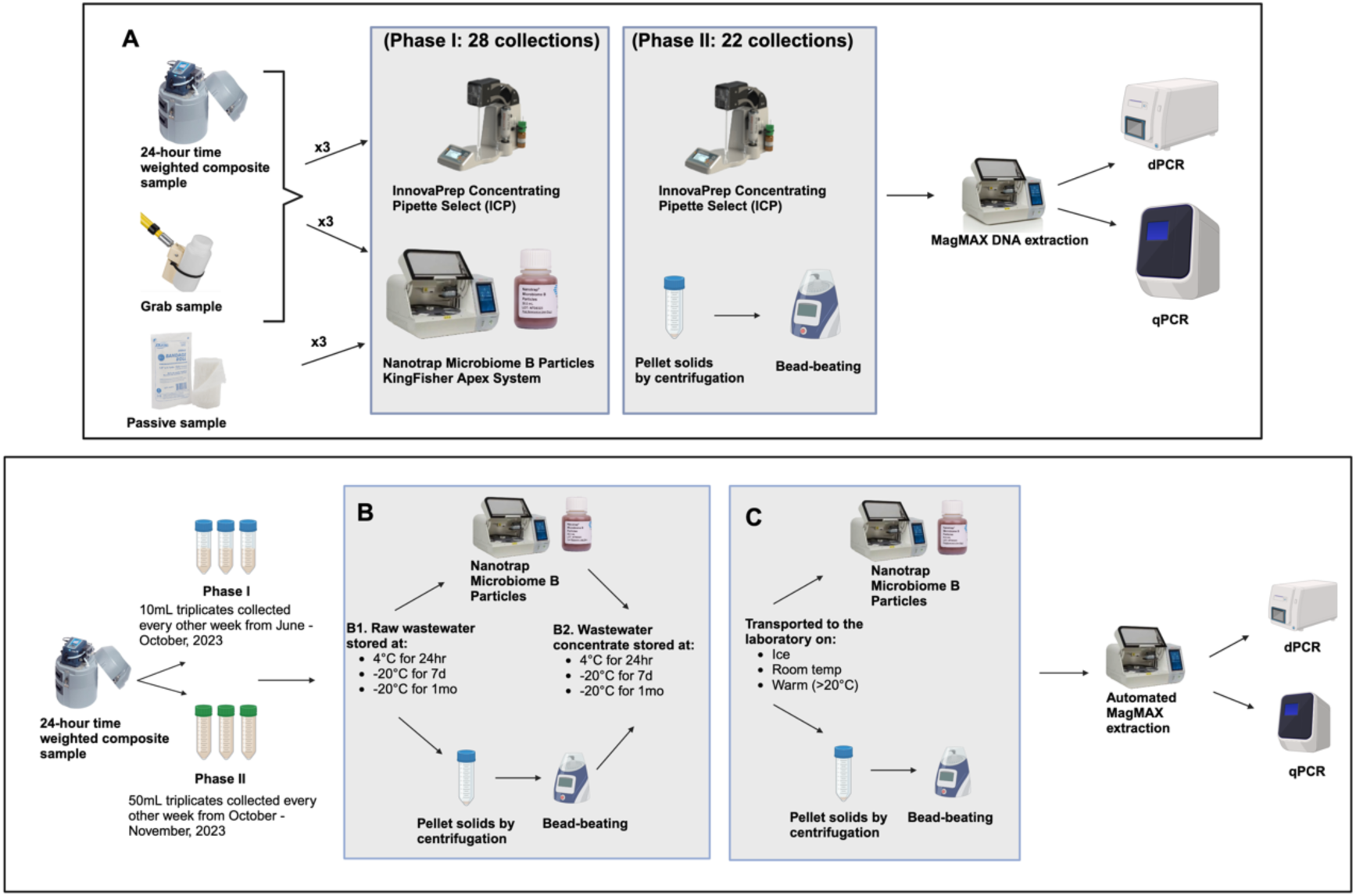
Experimental design. The regular collections (A) included comparisons of collection, concentration, and PCR methods. Composite and grab samples were initially concentrated with two different methods while passive samples were only concentrated with magnetic nanoparticles due to the limited volume obtained from Moore swabs. Halfway through the study (collection 29), we switched from magnetic nanoparticles to centrifugation and bead-beating to improve fungal DNA yield. All samples were extracted with the same method and resulting DNA was assayed by both qPCR and dPCR. Special collection compared sample storage (B) and transportation (C) conditions.

### Sample transport and storage experiments

Additional wastewater collections were carried out in triplicate once every other week for ten collections to test sample transportation and storage conditions (Fig. 1B). For the transportation experiments, samples were delivered to the lab (∼15 min. away) 1) on ice; 2) at room temperature (average 24°C); and 3) on heating pads (average 30°C).

For the storage experiments, both raw wastewater and wastewater concentrate (see below) samples were either processed immediately upon arrival or maintained at 4°C for 24 h; - 20°C for 7 days; or-20°C for 1 month prior to processing and analysis.

### Concentration

We initially compared two different concentration methods in triplicate: magnetic bead-based concentration using Microbiome B Nanotrap particles + Enhancement Reagent 3 (Ceres Nanosciences, Manassas, VA) and a filtration-based method using the Concentrating Pipette Select (InnovaPrep, Drexel, MO) (Fig. 1A, supplemental material). *Clavispora lusitaniae* was added as a process control to all samples prior to concentration (strains and details in supplemental materials). Because we observed lower than expected *C. auris* and *C. lusitaniae* DNA yields for wastewater collections 1-28 (Fig. 2), we modified our concentration method to replace Nanotrap concentration with concentration by centrifugation followed by bead-beating (supplemental materials) for wastewater collections 29-50.

**Figure 2.**
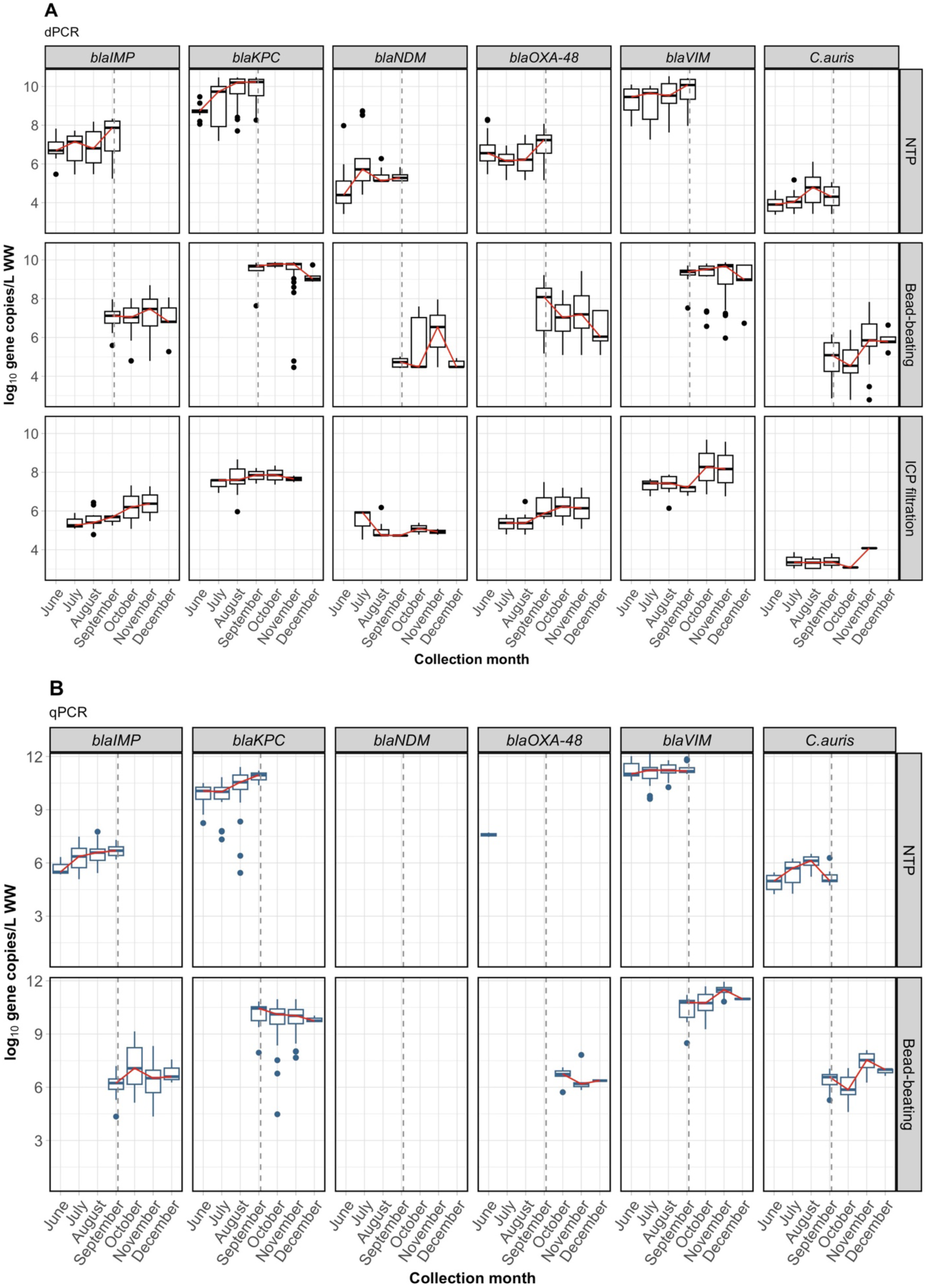
The distribution of log10 gene copies/L of wastewater for six monitored gene targets across months (x-axis) as determined by dPCR (A) or qPCR (B), with non-detects occuring with detects in replicate samples excluded. Preliminary analysis indicated that treating non-detects as ‘true’ zeros significantly inflated variance across replicates. Each facet corresponds to a gene target assayed. For dPCR, data is shown for passive sample replicates (n=58) derived from 22 collections, grab sample replicates (n=42) from 19 collections, and composite sample replicates (n=44) from 21 collections. For qPCR, only passive replicates (n = 145) obtained from 49 collections are included. The dashed gray line represents the concentration method transition from Nanotrap to bead-beating and centrifugation.

### DNA extraction and PCR analysis

Following concentration, nucleic acids were extracted using MagMAX Viral/Pathogen Nucleic Acid Isolation reagents on the Kingfisher Apex System (ThermoFisher, Waltham, MA) (supplemental materials).

We quantified *C. auris,* carbapenemase genes, and *Carjivirus communis* (formerly crAssphage, a human gut bacteriophage used as an endogenous control) using two different molecular assays: 1) qPCR with previously developed CDC assays and 2) dPCR with commercially available assays (GT Molecular, Fort Collins, CO) developed in contract with CDC’s National Wastewater Surveillance System and optimized for dPCR (GT-Digital AMR and DNA Pathogen Wastewater Surveillance Panel v1.0 and GT-Digital *C. auris* Wastewater Surveillance Assay Kit for the QIAGEN QIAcuity^®^ Digital PCR System v2.0) according to manufacturer’s specifications (supplemental materials).

CDC primers were used for *bla_KPC_* [12], *bla_NDM_* [12, 13], *bla_VIM_,* and *bla_OXA-48-like_* [14] and from previous studies for *bla_IMP_* [15], *C. auris* [16], and *C. communis* [17] for qPCR on a QuantStudio5 384-well instrument (supplemental materials).

### Cultivation of CPOs and *C. auris*

Select composite samples (50 ml) were pelleted at 4,500*xg* for 10 minutes and cultured for CPOs according to [18] [19] and *C. auris* according to [20] and CDC protocol FRL-100-P03 [12] (supplemental materials). Unique colony morphologies consistent with CPOs or *C. auris* were identified to species level using MALDI-TOF mass spectroscopy (bioMérieux, Marcy-I’Étoile, France). Presumptive CPOs were tested for carbapenemase genes using the Xpert^®^ Carba-R system (Cepheid; Sunnyvale, CA). All cultivation was done in triplicate with appropriate controls.

### Shotgun Metagenomics

Metagenomic sequencing was performed on composite wastewater, enrichment broth cultures, and selective plating cultures (supplemental materials). Microbial DNA was extracted using a QIAamp PowerFecal Pro DNA Kit (Qiagen), and libraries were prepared using a Nextera XT DNA Library Preparation Kit (Illumina). Pooled libraries were sequenced on a NovaSeq X instrument, employing paired-end 2×150 base reads (Accession number XXXXXX). Data analysis is described in the supplemental methods.

### Statistical analysis

All analyses employed multilevel models to account for repeated measurements within collection date and within triplicates (as appropriate). Quantitative detection level was log-transformed (base 10) for all biomarkers except *C. lusitaniae,* for which percent recovery was modeled without transformation. We primarily considered three indicators of replicate detection and variability in level of detection: 1) likelihood of detection (vs non-detection), 2) count of number of detections within three replicates (range 0-3), and 3) variance in quantitative detection level within triplicates (excluding non-detection). For detection models, we employed 3-level (detection within triplicate within collection date) logistic models to assess the odds of detection as a function of collection method, processing method, collection conditions, and diagnostic assay.

For count and variance models, we used 2-level (count or variance with collection date) linear models testing outcomes (count or variance) as a function of collection method, processing method, collection conditions, and diagnostic assay. Preliminary analyses showed that in the presence of non-zero values within triplicates, zeros were likely “false” zeros. Further, treating non-detects as “true” zeros led to artifactual inflation of the variance across triplicates. By removing non-detects for count and variance models only, we assessed the amount of variation in level for those instances with 2-3 positive detections.

A fourth set of analyses modeled quantitative level of detection as the outcome. For these models, we employed 3-level (level of detection within triplicate within collection date) linear models to assess levels of the outcome as a function of collection method, processing method, collection conditions, and diagnostic assay. All analyses were performed with SAS version 9.4 (Cary, NC).

## Results

### Comparison of dPCR and qPCR

All 6 molecular targets were detected in the facility wastewater by dPCR (*C. auris*: 370/750 (49%) samples including all replicates; *bla_KPC_:* 747/747 (100%); *bla_NDM_*: 362/748 (48%); *bla_VIM_*: 740/748 (99%); *bla_IMP_*: 709/748 (95%); and *bla_OXA-48-like_*: 638/748 (85%)), although with varying reproducibility between replicate samples (Fig. S2, Fig. 2). *C. auris* showed slightly lower gene copy numbers and lower odds of detection by dPCR (370/750 [49%] samples) compared to qPCR (411/747 [55%] samples) (odds ratio [OR] = 0.65, 95% confidence interval [CI] 0.46 – 0.93; Fig. 2, Table S3) The variance within triplicates was significantly higher for dPCR compared to qPCR (p = 0.002) (Fig. S2, Table S3) while the intraclass correlation for dPCR (0.87) and qPCR (0.96) suggested that both approaches had minimal residual variance within triplicates for *C. auris*.

By contrast, dPCR was more sensitive while qPCR was unable to reliably detect *bla_NDM_* (362/748 [48%] of dPCR samples vs. 38/748 [5%] of qPCR samples, OR = 37.1) and *bla_OXA-48-like_* (638/748 [85%] of dPCR samples vs. 15/149 [10%] qPCR samples, OR = 2958.3; Fig. 2, Table S3, Fig. S2). The most abundant carbapenemase gene, *bla_KPC_*, was detected in all samples with no significant different within-triplicate variance for dPCR vs. qPCR (p=0.6, intraclass correlation coefficient: dPCR= 0.95 and qPCR= 0.97). The variances within triplicates for *bla_IMP_* and *bla_OXA-48-like_* were significantly higher (p= 0.002 and 0.019, respectively) for qPCR compared to dPCR. Detections of *bla_VIM_* and *C. lusitaniae* were not significantly different for any of the models between qPCR and dPCR (Figs. 2, S3, Table S3).

### Comparison of grab, passive, and autosampler collections

Overall, the impact of the collection method was minimal (Table S4). Only *bla_VIM_* and *bla_OXA-48_* showed significant within-triplicate variance by collection method, with more variance in passive samples (*bla_VIM_*; p= 0.024) compared to autosampler composites and more variance in composite (*bla_OXA-48_*; p=0.006) compared to passive samples. There were minor differences in the number of detections within triplicates, varying by target of interest. *C. auris* averaged more detections within triplicate for passive compared to composite samples, (p= 0.027; Fig. 3, Table S4); *C. communis* averaged slightly fewer detections with passive samples compared to composites, (Fig, S4, Table S4); *C. lusitaniae*, averaged slightly more detections with grab samples compared to composites, (p= 0.044; Table S4); and *bla_OXA-48_* showed fewer detections in grab samples compared to composites (p= 0.004; Table S4). The mean *C. lusitaniae* recovery was highest for passive samples (378%), followed by composite (237%) and then grab samples (177%).

**Figure 3.**
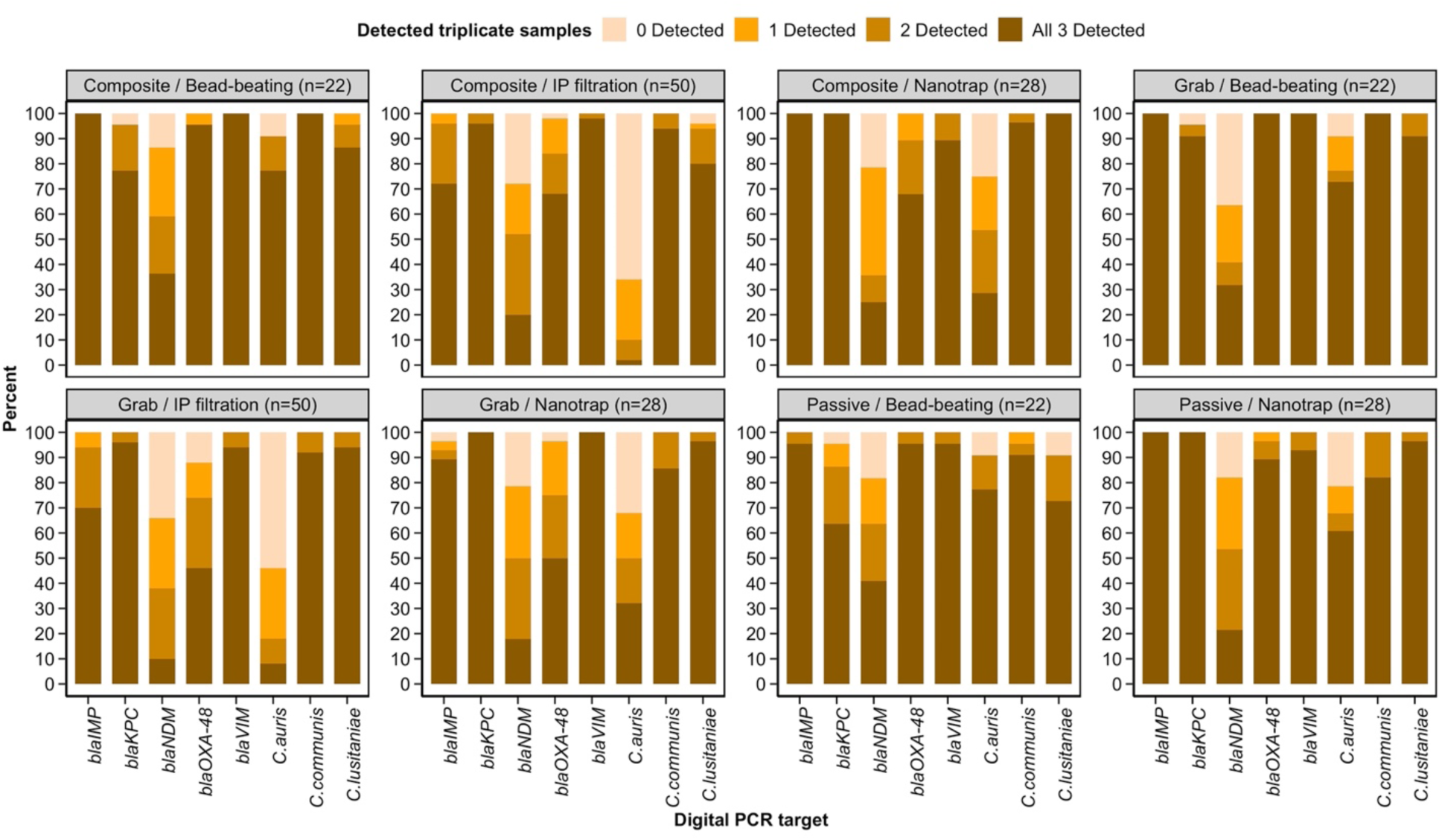
Collection and concentration methods by number of replicates detected by digital PCR. Bar plots illustrate the percentage of triplicates detected for eight biomarkers (labeled along the x-axis) across a detection scale ranging from 0 to 3. Detection levels are represented by the colors, while each facet represents a distinct collection and concentration method. The total number of collections included in each facet is indicated, with each collection processed in triplicate.

We investigated the impact wastewater parameters (i.e., flow rate, temperature, turbidity, and pH), most of which were consistent over time, although periodic large flushes of water were observed in manhole (Fig S5). Across all models predicting detection and variance within triplicates, the effects of wastewater collection parameters were generally non-significant (not shown). For multivariate models predicting gene copy numbers using wastewater data parameters, several significant effects of emerged: higher flow rate and temperature were associated with higher levels of detection for *C. auris (*p *<*0.001); greater turbidity was associated with higher levels of detection for *bla_KPC_* (p-0.008); and higher pH was associated with higher recovery of *C. lusitaniae* (p=0.02).

### Comparison of wastewater concentration methods

Because of the change in concentration methods, we conducted models separately for the first and second halves of the sampling period, controlling for collection method (Table S5).

Compared to ICP, NP yielded higher average number of within-triplicate detections for *C. auris* (p< 0.001), *C. lusitaniae* (p= 0.003), and *bla_IMP_* (p= 0.009) (Fig. 3, Table S5). With bead-beating, *C. auris, bla_NDM_*, *bla_IMP_*, and *bla_OXA-48-like_* detections and gene levels increased significantly (p< 0.001 for all) relative to the ICP concentration (Figs. 2B, 3, Table S5). Though there were significant differences in within-triplicate variation for both *bla_KPC_* (p=0.003) and *C. communis* (p= 0.003) (favoring ICP over NP), and for *bla_VIM_* (p< 0.001), *bla_IMP_* (p= 0.002), and *bla_OXA-48-like_* (p= 0.001) (favoring bead-beating over ICP), the differences were modest (Fig 3, Table S5).

### Effects of sample transport and storage conditions

There was no significant impact on detection or within-triplicate variance (Table S6) with different transportation temperatures (on ice, 25°C, and >25°C). There were, however, significant increases in the detection levels of *bla_KPC_* (p= 0.003) and *bla_VIM_* (p<0.001) when samples were transported at room temperature and of *C. communis* (p=0.024), *bla_KPC_*, (p< 0.001), *bla_VIM_* (p< 0.004), and *bla_OXA-48_* (p= 0.024) when samples were transported warm (Table S6), compared to transportation on ice.

Models predicting detection vs. non-detection showed no significant effects based on storage conditions for any of the targets except *bla_NDM_*, which had a significantly higher likelihood of detection in raw samples stored at-20°C for 7 days and 1 month compared to no storage (OR=4.2, p=0.03 and OD=14.8, p<0.001, respectively; Table S7). For *C. auris*, each of the 6 storage conditions had lower variance than the no storage condition (*P* between 0.006-0.051). Finally, for models predicting the level of detection, there were no significant effects on gene copy numbers for *bla_KPC_, C. communis*, and *C. lusitaniae,* but for *C. auris*, *bla_NDM_, bla_VIM_, bla_IMP_*, and *bla_OXA-48-like_*, there were inconsistent patterns of effects (Table S7). For example, for raw samples stored at any temperature/duration, the levels of *C. auris* detection were lower (*P* values from 0.01 to 0.06) than immediate processing but for concentrate stored at any temperature/duration, the levels of detection were higher (*P* values from 0.03 to 0.05) than immediate processing.

### Identification of CPO taxa and *C. auris* via cultivation and metagenomic sequencing

Although there were PCR detections of several targets for which no cultured representatives were obtained (e.g., *C. auris* was cultured from 9/10 of wastewater samples (Fig. 4) but was detected by PCR in all 10), no genes were culture-positive but PCR negative. The most common carbapenemase-producing organisms recovered in culture were *Aeromonas* and *Citrobacter* species with KPC genes and *Pseudomonas putida* with VIM genes (Fig. 4), representing organisms not commonly recovered from patients in either in colonization or infection state [21]. In contrast, putative patient-colonizing MDROs such as KPC-producing *Klebsiella pneumoniae* and NDM-producing *E. coli* were less frequently isolated from the wastewater samples (Fig. 4).

**Figure 4.**
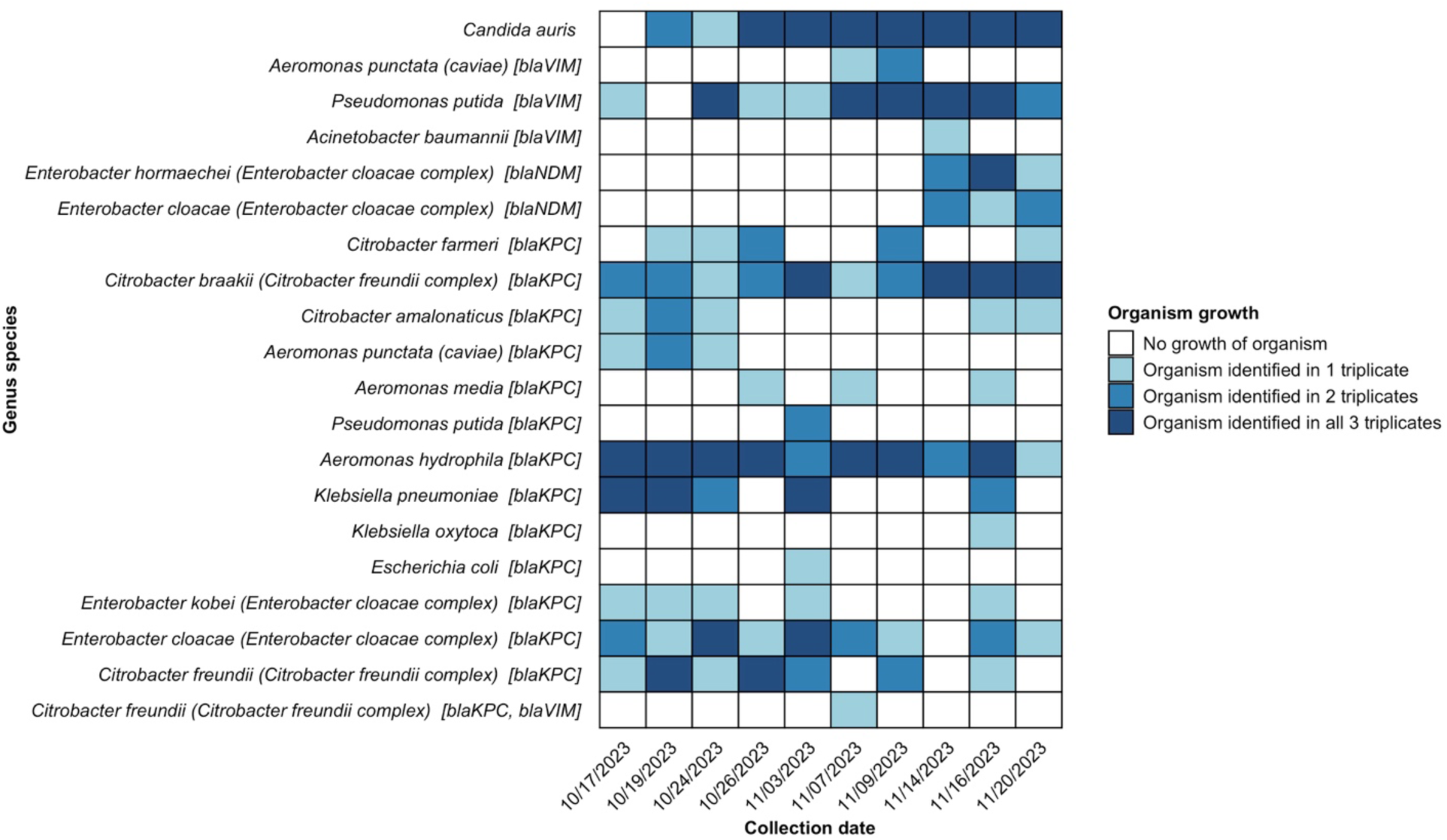
Carbapenemase-producing organisms recovered from ten composite samples (cultured in triplicate) spanning from October 12, 2023 to November 20, 2023. The heatmap illustrates the presence of organisms in cultured triplicates, with a color gradient indicating the levels of growth from no organism detected to organisms identified in all triplicates. Text in the brackets next to the y-axis labels denote the specific carbapenemase genes associated with the isolated gram-negative bacteria.

Metagenomic sequence data from wastewater-derived samples supplemented culture results (Fig. 5). Only a small percentage of the total antimicrobial resistance genes identified in the metagenomes from raw wastewater (4.5%), broth (6.2%), and plates (7.8%) fell within the carbapenemase gene families quantified by PCR. Among those, there were 8 unique carbapenem resistance gene alleles identified across sample types including several previously reported as human clinical pathogens (VIM-2 [22], VIM-4 [23], VIM-12 [24], KPC-2 [25], and KPC-3 [26]) and some which have not (KPC-25, KPC-30, and KPC-34). The PCR primers/probes used here aligned with no mismatches to all alleles detected in metagenomes, indicating that the carbapenemase genes in the metagenomic datasets were likely to have been quantified by PCR. Three of the 49 metagenomic datasets (6%) had no hits to genes found via PCR. There were no detections of genes producing IMP, OXA-48, or NDM in the metagenomes despite detection by dPCR (93% of samples had *bla_IMP_*, 92% *bla_OXA-48_*, 57% *bla_NDM_*). Although it is difficult to determine which organisms harbored the AR genes identified in the metagenomes and nearly 40% of sequences could not be taxonomically classified, the most abundant identified members of the metagenomes, e.g., *Aeromonas* (7%)*, Acinetobacter* (12%)*, Enterococcus* (4%)*, Klebsiella* (3%), and *Pseudomonas* (18%) were also among the cultured isolates (Fig S6).

**Figure 5.**
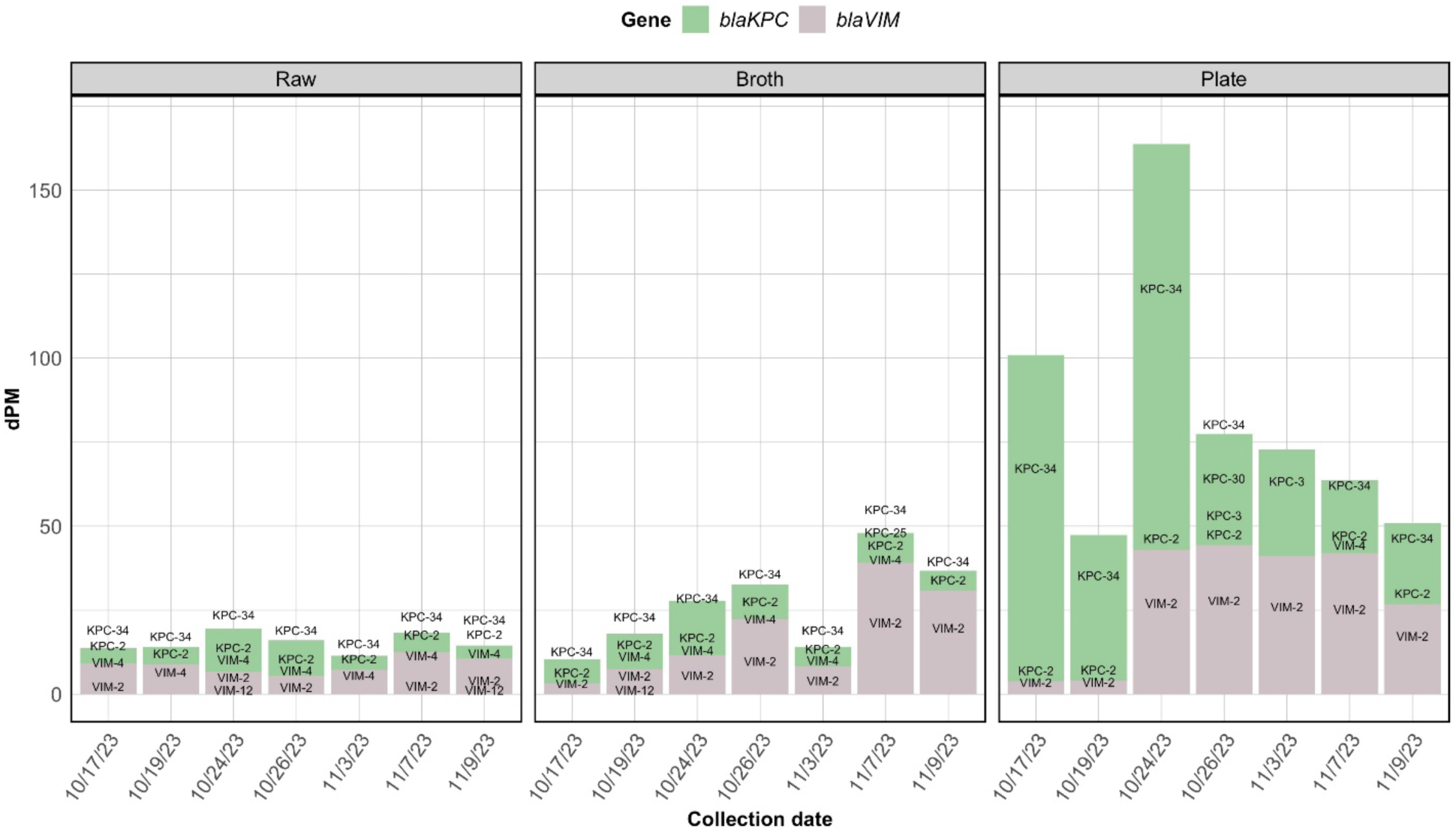
Relative abundance (in dPM) of queried carbapenem resistance gene alleles in raw wastewater (Raw), enriched broth cultures (Broth), and selective plating cultures (Plate). Mean relative abundance metric reads depth per million (dPMs) for each allele in Broth and Plate triplicates are shown for clarity. The labels on the plot are individual gene alleles for the resistance genes depicted.

## Discussion

### Wastewater sampling and assay methods

Procuring wastewater samples from healthcare facilities may be one of the biggest challenges for surveillance efforts due to facility willingness, service access logistics, and adequate water flow. We found that once an appropriate access point was identified, different collection methods were similar with respect to reliability and detection. Passive and composite samples are generally thought to be more representative than grab samples which represent a snapshot of a single point in time [27]. Grab and composite samples have been shown to correlate well for SARS-CoV-2 virus [28] with improvements in detection from passive samples relative to grab [29]. Here, we found passive and grab sampling to be more logistically reliable and less costly compared to autosamplers, with passive samples yielding a higher recovery of the spiked-in control. We therefore recommend passive sampling when feasible. Our observation of increased fungal detection during high flow conditions may not be biologically relevant, but could indicate the dilution of inhibitors in the wastewater or the introduction of compounds that could contribute to the lysis of fungal cells; we therefore recommend monitoring flow rate during sample collection. Although we observed few differences in downstream detection whether samples were transported to the lab on ice, at room temperature, or warm, we recommend rapid processing of samples or, at minimum, consistent storage methods. That said, this study was limited by a relatively small sample size of 50 regular collections and 10 each of the transport and storage collections, so non-significant findings do not necessarily mean that some of the methods or conditions do not impact detection.

The *C. lusitaniae* recoveries were often >100% indicating an underestimate of the spike-in quantity. While this may result in imprecise measurements, it is sufficient for calculating recovery and comparing results within this study to evaluate the performance and reliability of the overall analytical workflow.

Once collected, WWS workflows employ a variety of concentration methods, including centrifugation, membrane filtration, chemical precipitation, ultrafiltration, and magnetic particles that can impact target detection in WWS. [4, 30]. We found that bead-beating is needed for *C. auris* detection by PCR and does not significantly impact other target detections, supporting prior observations of improved *C. auris* detection with bead-beating to lyse cells [31]. Here, we coupled bead-beating with concentration by centrifugation, although it is possible to use NTP concentration instead [12].

PCR comparisons highlighted the challenges of quantifying genetic targets in wastewater compared to clinical samples. Detection of AR targets across three replicates showed variable reliability, reflecting the heterogeneity of the wastewater matrix. Significant variability can impair accurate quantification of AR targets, underscoring the importance of performing quantification in replicate and establishing baseline measurements as a practical solution.

dPCR assays using commercially-available kits designed in partnership with CDC’s National Wastewater Surveillance System out-performed qPCR assays for *bla_IMP_, bla_NDM_, bla_VIM_,* and *bla_OXA-48-like_*. The primer sequences were the same for both qPCR and dPCR for all genes but *bla_IMP_,* which was designed using a set of reference genes from the NCBI MicroBIGG-E database and covers many but likely not all *bla_IMP_* genes (personal communication, GT Molecular); qPCR primers involved separate oligonucleotides for *bla_IMP-14_* and *bla_IMP-4_* only. The probe sequences for several of the dPCR assays were modified from the qPCR sequences to increase the melting temperatures (Tm) for qPCR probe oligonucleotides. We suspect that the Tm modification accounts for the improvement in the dPCR assays over qPCR and that the qPCR non-detection of carbapenemase targets like *bla_NDM_* likely represent false negatives.

Nevertheless, the higher sensitivity we observed with dPCR for *bla_NDM_* and *bla_OXA-48-like_* is supported by previous studies of carbapenemase [32] and SARS-CoV-2 genes [33] in wastewater. Interestingly, qPCR outperformed dPCR for the fungal targets examined here, though other studies have shown the opposite for fungal detection in clinical samples (e.g., [34–36]). We attempted to determine if this was due to co-located gene copies of the fungal ribosomal RNA operon (data not shown). The *C. auris* chromosome 5 contains 9 total copies of ITS2; multiple copies in a single dPCR well could be counted as a single detect vs. multiple qPCR detects. Tests with increased acoustic DNA shearing reduced the discrepancy between dPCR and qPCR, but not enough to account for the difference (data not shown).

### Clinical relevance of wastewater data

Although the goal of establishing WWS is to detect patient shedding in healthcare facilities, this study did not incorporate longitudinal sampling of patients (e.g., serial point prevalence surveys). Our foundational methods and baseline target assessment proved important given the detection of potentially background CPOs in wastewater. Carbapenemase genes are frequently encoded by genes located on mobile genetic elements, facilitating their exchange and dissemination [37]. Although clinically relevant *bla_KPC_*, *bla*_NDM_, and *bla*_OXA-48_ have been identified in hospital and municipal wastewater [38], both sewers [39] and hospital wastewater [7, 40] are also known to be inhabited by diverse microorganisms, not all of fecal origin.

Furthermore, AR genes can often be found in these organisms [7], which could serve as reservoirs/vectors for environmental spread of carbapenemase genes.

We used culture-and sequence-based approaches to evaluate whether the genes detected by qPCR and dPCR were clinically relevant. Preliminary culture data on a limited number of samples (n=10) revealed *C. auris* and a large reservoir of carbapenemase-producing organisms inhabiting the wastewater ecosystem, potentially independent of patient organism burden.

Longitudinal surveillance of healthcare facility wastewater should therefore include organism identification (including non-Enterobacterales), establishment of baseline gene levels, or characterization of wastewater-origin AR burden (within water or in biofilms). A better understanding of the evolution and spread of *C, auris* and AR from patient/hospital to sewer will also help in the interpretation of WWS data.

While dPCR can quantify gene abundance, metagenomics provides an additional layer of allele-specific relative abundances. Three of the alleles that we identified by metagenomics have not been previously associated with humans and may represent colonization of the plumbing system or undetected human colonization. However, dPCR is more practical for routine surveillance of carbapenemase genes and may be more sensitive at low gene abundances. For example, there were no detections of *bla_IMP_*, *bla_OXA-48_*, or *bla_NDM_* by non-targeted metagenomic sequencing though these genes were detected by targeted dPCR assays. dPCR also appeared to capture all alleles detected by metagenomics. Periodic metagenomic monitoring can confirm if dPCR assays are detecting dominant carbapenemase genes and identify novel targets.

We show that WWS of *C. auris* and CPOs is possible in a healthcare setting and provide guidance on best practices (summarized in Table 1). Evaluating clinical relevance will require further monitoring and include correlations with patient pathogen prevalence.

**Table 1:**
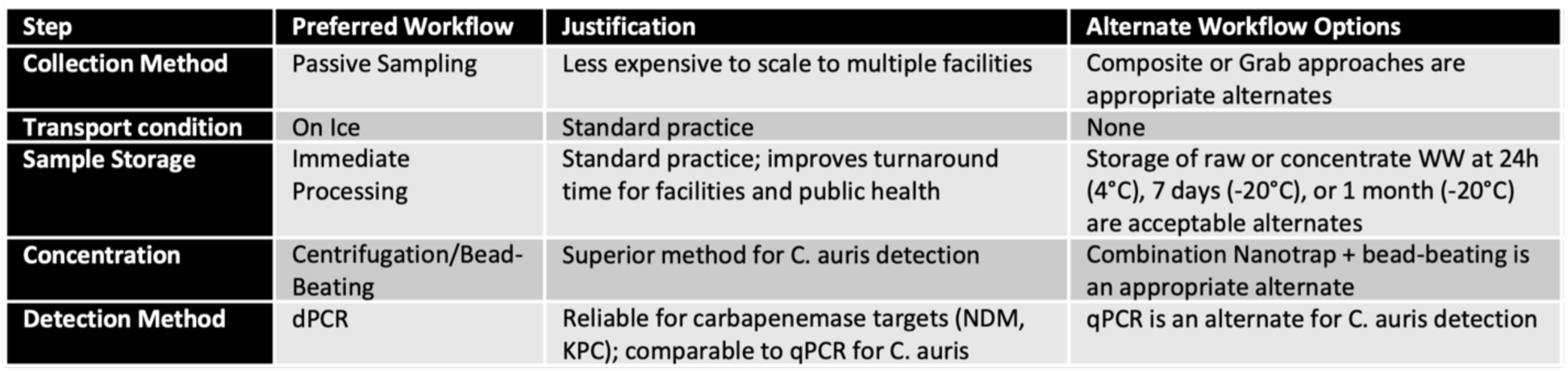
Recommended Workflow for Wastewater Surveillance.

## Data Availability

All data produced in the present study are available upon reasonable request to the authors

## Acknowledgements

We gratefully acknowledge the time and effort of our partners at Rush University Medical Center: Pamela Bell, Jeremy Kahsen, McKenzi King, Kevin Kunstman, Siri Pothula, Sonia Sherwani, Lahari Thotapalli, and Robert Weinstein; Chicago Department of Public Health: Kendall Anderson, Regina Atwater, Stephanie Black, Peter Dejonge, Dorothy Foulkes, Richard Teran, Do Young Kim, Alyse Kittner, Colin Korban, Hira Adil, Massimo Pacilli, Haifa Wahbeh, Kelly Walblay, Christy Zelinski; LTACH staff: Lisa Duffner, Frederick Nartey, Nidhi Undevia, Patrick Geary; GT Molecular: Sarah Kane, Audrey McDonald, Caleb Willis, Max Zvyagin.

Quantitative PCR, digital PCR and metagenome sequencing were performed by members of the Genomics and Microbiome Core Facility (GMCF) at Rush University.

## Author contributions

DSC and AH led the field and lab work and wrote the manuscript with RP; C-YL, MJL, MS, CC, and EG conducted lab and field work; EN and AC generated and analyzed sequence data; AC-S, AL, and FW helped conceive of and supervised the work; MS conducted statistical analyses; LD and NU provided facility access and data; SG, MH, and ML, and RP acquired funding, supervised the work. All authors reviewed and edited the manuscript.

