## Supplemental materials for "Sewer monitoring for antimicrobial resistance genes and organisms at healthcare facilities"

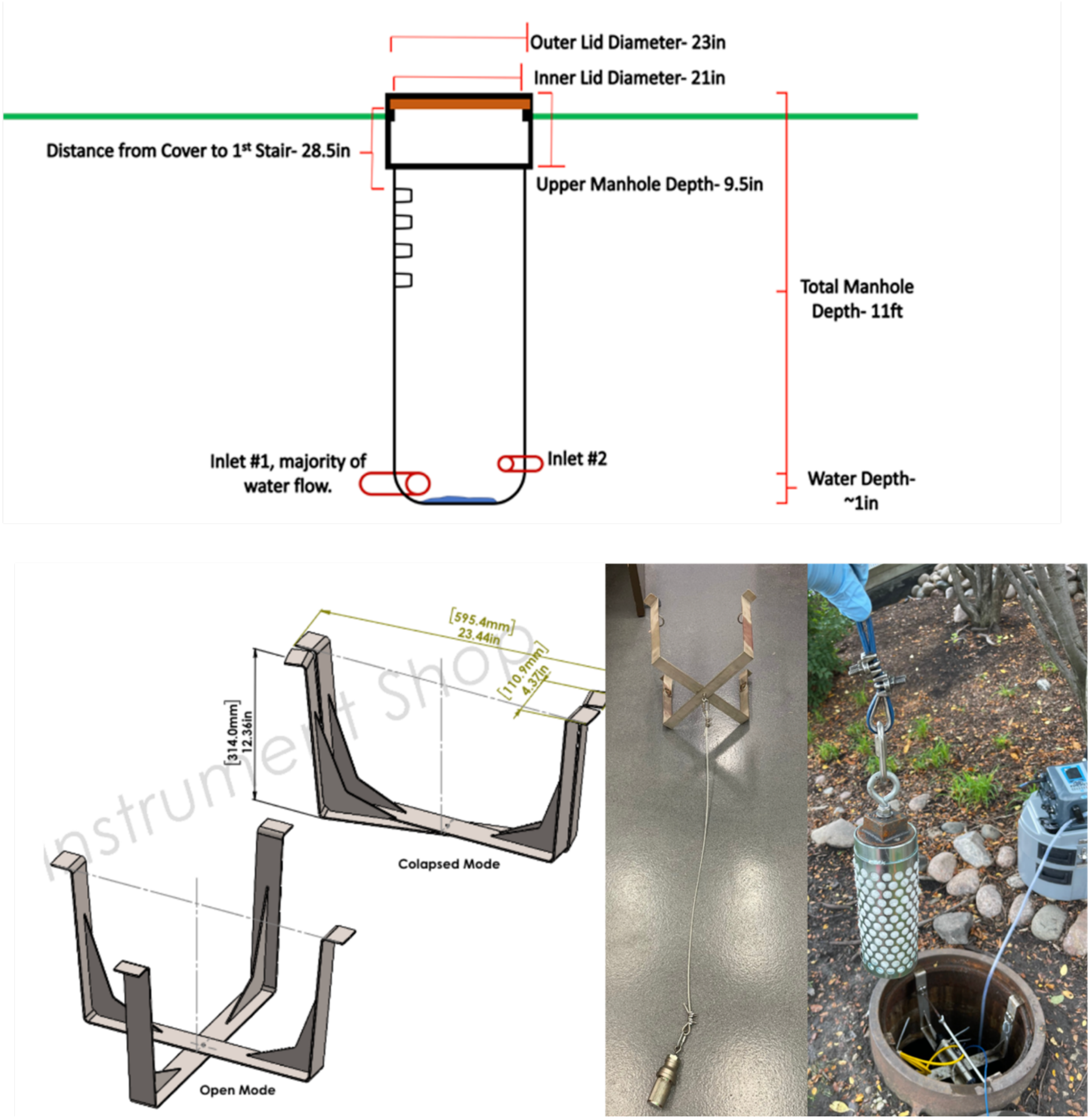

Figure S1. Schematic of the manhole (top), the rig bracket manufactured in the UIC instrument shop (bottom left), and photos of the rig with attached cable and cannister in the lab and in use (bottom right).

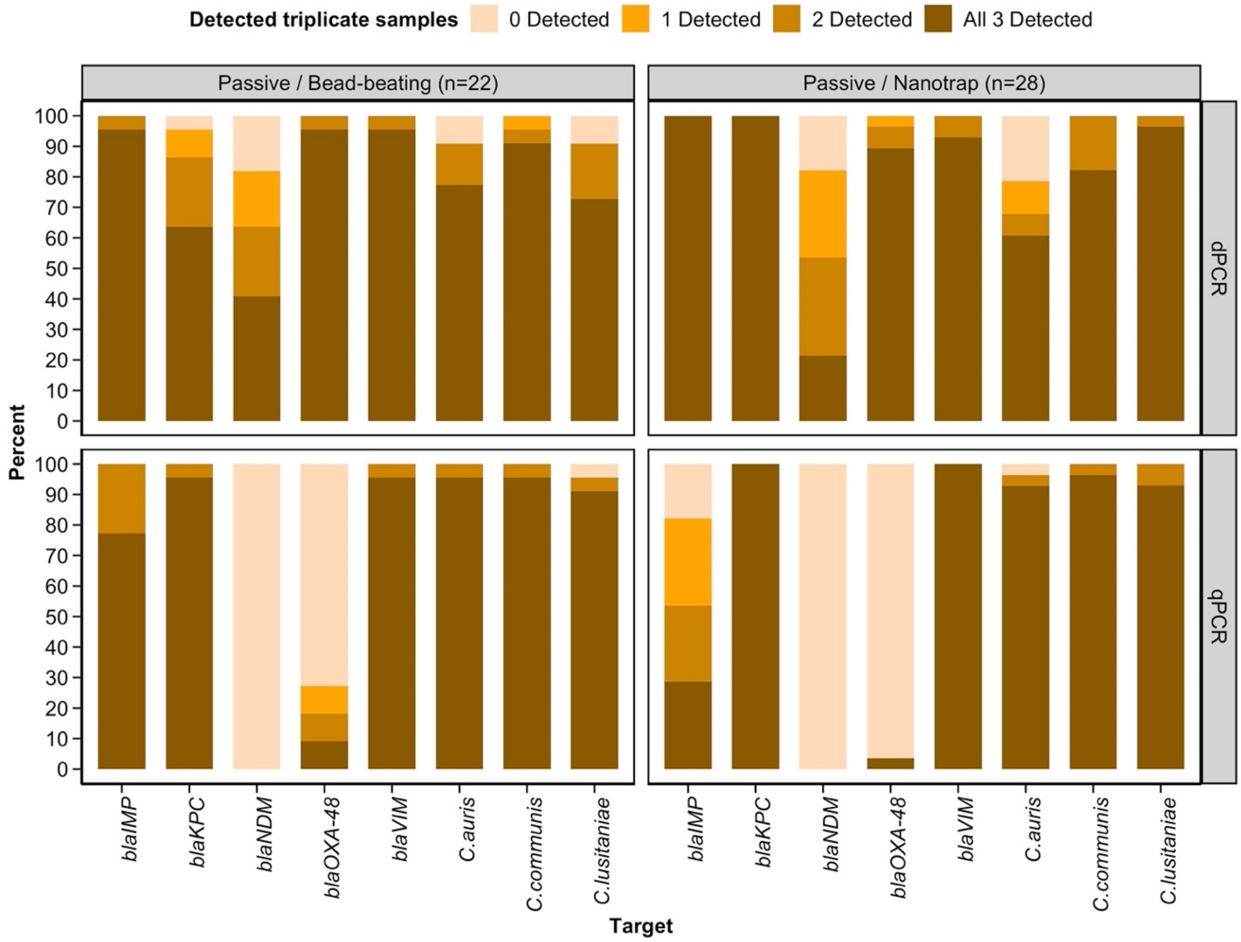

Figure S2. Bar plots illustrating the percentage of triplicate detections for the eight gene targets (labeled across the x-axis) using the passive collection method. Detection levels are represented on a scale from 0 to 3, with each level indicated by distinct colors. The vertical facets correspond to the two PCR quantification techniques employed in this study, while the horizontal facet indicates the collection and concentration method with the total number of collections analyzed indicated.

.

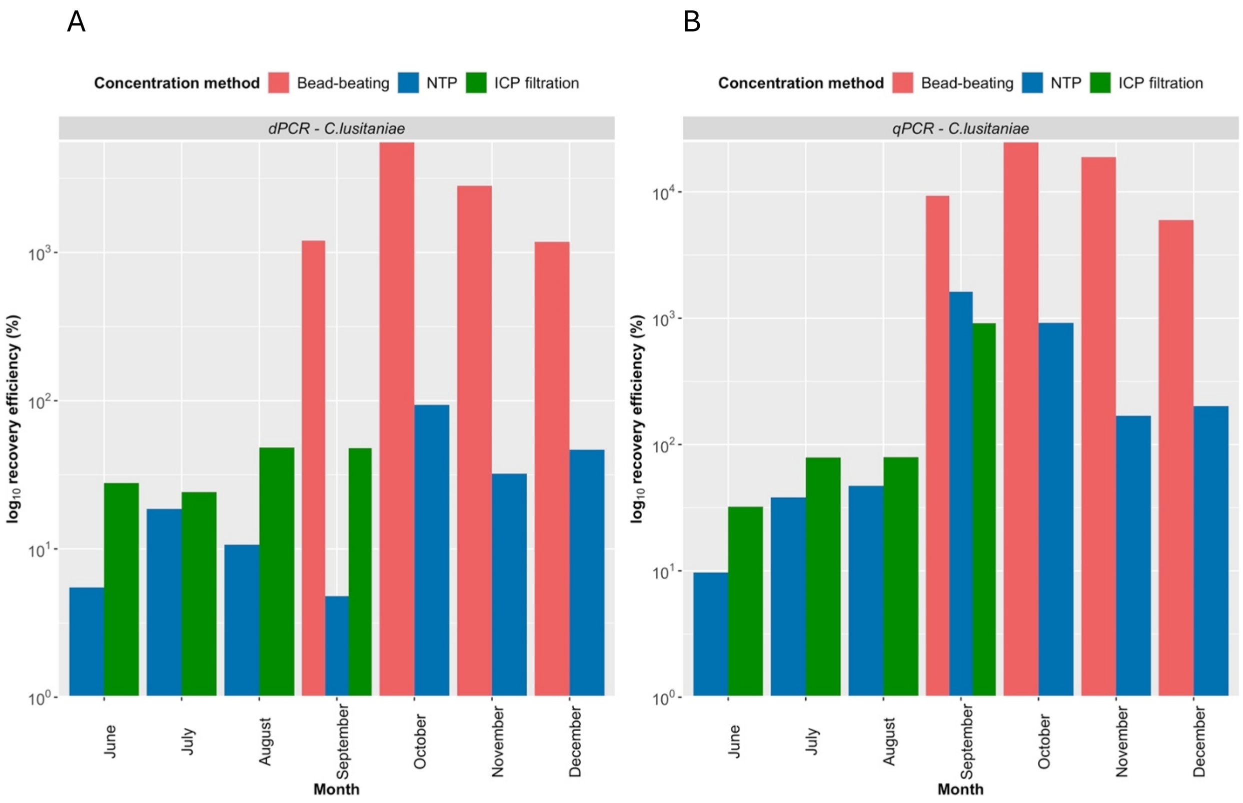

Figure. S3. Bar plots illustrating the recovery efficiency of *C. lusitaniae* as the process control for samples analyzed over a six-month period. Colors indicate the different concentration methods used. The facets correspond to the biomarker, with panel sections A and B denoting the results from the PCR quantification methods from the analysis.

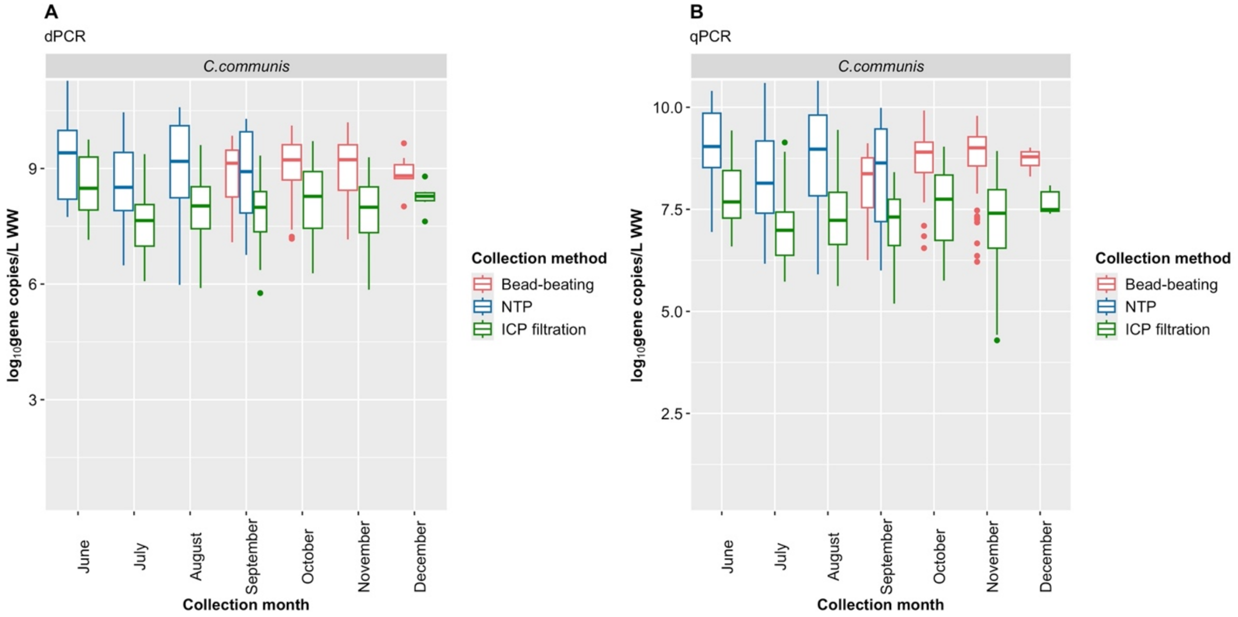

Figure. S4. Box plots illustrating the distribution of log10 gene copies/L of wastewater for fecal indicator, *Carjivirus communis* as determined by dPCR (A) and qPCR (B), with non-detects excluded. The colors of each box plot indicate the concentration method used (Centrifugation + bead-beating, Nanotrap particles, Innovaprep Concentrating Pipette [ICP]).

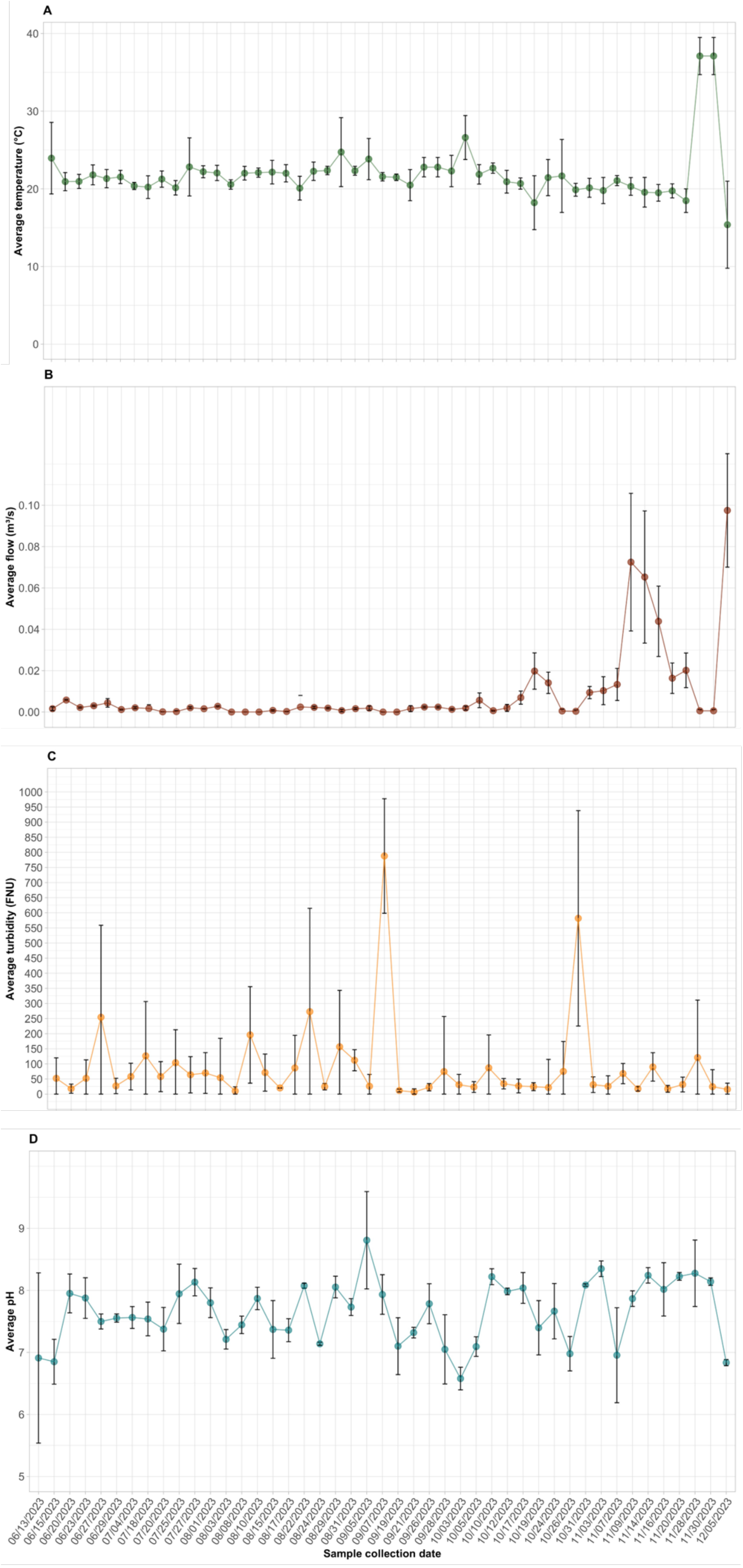
A
Average temperature (°C)
B
Average flow (m³/s)
C
Average turbidity (FNU)
D
Average pH
06/13/2023
06/15/2023
06/20/2023
06/23/2023
06/27/2023
06/29/2023
07/04/2023
07/18/2023
07/20/2023
07/25/2023
07/27/2023
08/01/2023
08/03/2023
08/08/2023
08/10/2023
08/15/2023
08/17/2023
08/22/2023
08/24/2023
08/29/2023
08/31/2023
09/05/2023
09/07/2023
09/19/2023
09/21/2023
09/26/2023
09/28/2023
10/03/2023
10/05/2023
10/10/2023
10/12/2023
10/17/2023
10/19/2023
10/24/2023
10/26/2023
10/31/2023
11/03/2023
11/07/2023
11/09/2023
11/14/2023
11/16/2023
11/20/2023
11/28/2023
11/30/2023
12/05/2023
Sample collection date

Figure S5. Measured wastewater temperature (A), flow (B), turbidity (C), and pH (D) during the course of the study. The temperature and flow average values were calculated from measurements taken hourly for 24h preceding sample collection. Error bars represent standard deviation. For parameters (C), data for four collections are missing (07/25/2023, 07/27/2023, 08/01/2023, 09/12/2023) and for (D), data for five collections are missing (07/06/2023, 7/11/2023, 07/13/2023, 09/14/2023) due to instrument malfunction.

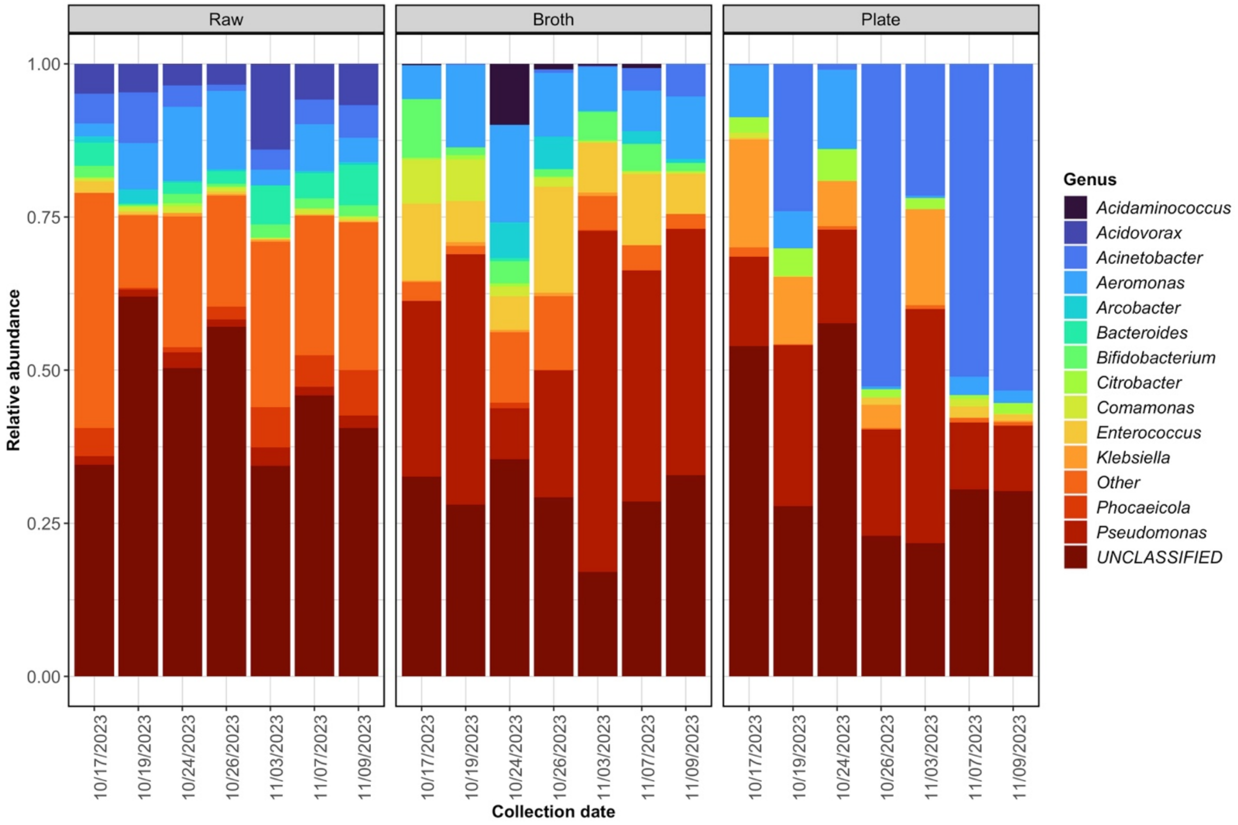

Figure S6. Relative abundance of different genera identified in metagenomes from raw wastewater (left), enrichment broth (middle), and isolation pates (right). The ‘other’ category includes organisms with <0.5% abundance in all samples. Unclassified reads could not be assigned to a taxon.

Table S1. Baseline information about LTACH A (June 2023)

| Covariate | Value |
|---|---|
| Census[a] | 37 |
| Licensed beds | 86 |
| Baseline prevalence of carbapenem resistant organisms targeted for this study (single day assessment on June 30, 2023), n (%) [b] | *1.* *C. auris*: 13 (35)<br>2. CRE: 13 (35)<br>3. CRAB: 3 (8) |
| Past 6 months of MDRO and CRO cases (January 1, 2023 to June 30, 2023), n (%) [b] | *1.* *C. auris*: 34 (18)<br>2. CRE: 19 (10)<br>3. CRAB: 9 (5) |
| Status of performing on-going/admission MDRO/CRO screening | No admission screening of MDROs or CROs |
| Number of patients using toilet or other waste management entering wastewater stream (excludes bed pad) | 14 / 37 patients (38%) based on entire month data ending June 30, 2023 data |

Notes.

a. Total staff and overall turnover rate were estimated from LTACH A Human Resources data from July 1, 2023 to August 31, 2023.

b. Baseline MDRO and CRO prevalence (single day assessment on June 30, 2023) and past 6 months of data (January – June 2023) were estimated by obtaining the census of patients, identifying those patients on Contact Precautions, and recording reason for Contact Precautions from the Electronic Medical Record. MDRO status may represent either past or current (presumed or confirmed) colonization status, which would lead to Contact Precautions. Carbapenem-resistant Pseudomonas aeruginosa infection-control information not available during study period, because it was not categorized in the electronic medical record for infection control. CRE = Carbapenem-resistant Enterobacterales. CRAB = Carbapenem-Resistant Acinetobacter baumannii.

**Table S2: Primer and probe sequences for qPCR assays**

| Assay Name | Organism Target / (Gene Target) | Oligo type | Primer Sequence | Reference |
|---|---|---|---|---|
| ***C. auris*** | *C. auris /* (ITS) | | | |
| | | Forward Primer | CAGACGTGAATCATCGAATCT | Leach et al. 2018 |
| | | Reverse Primer | TTTCGTGCAAGCTGTAATTT | |
| | | Probe | 5′-/56-carboxyfluorescein (FAM)/AATCTTCGC/ZEN/GGTGGCGTTGCATTCA/3IABkFQ/-3′ | |
| **Inhibition control** | *Drosophila melanogaster* / (Bicoid) | | | |
| | | Forward Primer | CAGCTTGCAGACTCTTAG | Leach et al. 2018 |
| | | Reverse Primer | GAATGACTCGCTGTAGTG | |
| | | Probe | 5′/Cy3/AACGCTTTGACTCCGTCACCCA/3IAbRQSp/-3′ | |
| **CPQ_056** | CrAssphage / ( 14731−14856 pos) | | | |
| | | Forward Primer | CAGAAGTACAAACTCCTAAAAAACGTAGAG | Stachler et al. 2018 |
| | | Reverse Primer | GATGACCAATAAACAAGCCATTAGC | |
| | | Probe | [FAM] AATAACGATTTACGTGATGTAAC [MGB] | |
| **Carbapenemase genes OXA-48-like and VIM** | VIM | Probe | FAM-TTA CCC GCA TCT ACC-BHQ1 | CDC ARC.TE.C.0143 |
| VIM - *Pseudomonas aeruginosa* AR Bank #0054 (pos) | OXA-48-like carbapenemases | Forward Primer | ACG GGC GAA CCA AGC AT | |
| OXA-48-like - *Klebsiella pneumoniae* AR Bank #0039 (pos) | Enterobacterales | Reverse Primer | GCG ATC AAG CTA TTG GGA ATT T | |
| OXA-48-like *Klebsiella pneumoniae* ATCC strain BAA-1706 (neg) | *Pseudomonas aeruginosa* | Probe | HEX-CGG AGA TTG ARaA AGC A-BHQ1 | |
| VIM *Klebsiella pneumoniae* ATCC strain BAA-1706 (neg) | *Acinetobacter baumannii* | Forward Primer | GAA AAA CAC AGC GGC ACT TCT | |
| | | Reverse Primer | CAC GCG TTA CRaG GAA GTC CAA | |
| **Carbapenemase gene KPC and NDM** | *Klebsiella pneumoniae* carbapenemase (KPC) | Probe | FAM-TG ATA ACG CCG CCG CCA ATT TGT-BHQ1 | CDC ARC.TE.C.0022 |
| KPC - Positive Control *K. pnuemoniae* ATCC BAA-1705 (pos) | New Delhi metallo-β-lactamase (NDM) | Forward Primer | GGC CGC CGT GCA ATA C | |
| NDM - Positive Control *K. pnuemoniae* ATCC BAA-2146 (pos) | | Reverse Primer | GCC GCC CAA CTC CTT CA | |
| BAA-1705 and BAA-2146 combination (50:50) (pos) | | Probe | HEX-TG GAT CAA GCA GGA GAT-BHQ1 | |
| KPC and NDM Negative Control – *K. pnuemoniae* ATCC BAA-1706 (neg) | | Forward Primer | GAC CGC CCA GAT CCT CAA | |
| | | Reverse Primer | CGC GAC CGG CAG GTT | |
| **Carbapenemase genes blaIMP** | Enterobacteriaceae (CRE) | | | |
| | IMP | Forward Primer | ATT TTC ATA GTG ACA GCA CGG GC | Pollett et al 2014 |
| | | Reverse Primer | CCT TAC CGT CTT TTT TAA GCA GCT CAT TAG | |
| | | Probe | HEX-TTC TCA ACT CAT CCC CAC GTA TGC-BHQ1 | |

**Table S3.** Comparison of (A) dPCR versus qPCR for outcomes of detection (yes/no), (B) number of detects (0-3), and (C) variance within triplicates with *C. lusitaniae* variance shown separately.

**3A. Logistics models predicting detection/non-detection**

| Target | Est | 95% CI lower | 95% CI upper | p-value |
|---|---|---|---|---|
| C. auris | -0.160 | -0.310 | -0.010 | 0.032* |
| blaKPC | 0.020 | -0.010 | 0.050 | 0.247 |
| blaNDM | 1.300 | 1.160 | 1.430 | < 0.001*** |
| C.communis | -0.030 | -0.060 | 0.000 | 0.075 |
| C.lusitaniae | 0.080 | -0.010 | 0.180 | 0.077 |
| blaVIM | -0.040 | -0.100 | 0.020 | 0.159 |
| blaOXA-48 | 2.600 | 2.360 | 2.840 | < 0.001*** |
| blaIMP | 0.840 | 0.560 | 1.120 | < 0.001*** |

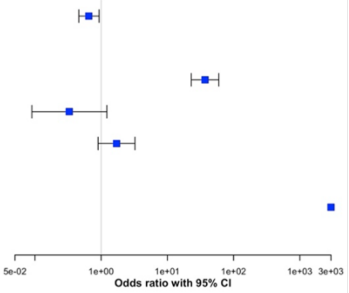

**Notes**
Results for blaVIM, blaOXA-48,and blaIMP account for only 1 collection method (passive) and 2 concentration methods (NTP and bead-beating)
-- = not applicable; all replicates had detects
OR = odds ratio (95% Confidence Interval)

**3B. Linear models predicting number of triplicates detected**

| Target | Est | 95% CI lower | 95% CI upper | p-value |
|---|---|---|---|---|
| C. auris | -0.160 | -0.310 | -0.010 | 0.032* |
| blaKPC | 0.020 | -0.010 | 0.050 | 0.247 |
| blaNDM | 1.300 | 1.160 | 1.430 | < 0.001*** |
| C.communis | -0.030 | -0.060 | 0.000 | 0.075 |
| C.lusitaniae | 0.080 | -0.010 | 0.180 | 0.077 |
| blaVIM | -0.040 | -0.100 | 0.020 | 0.159 |
| blaOXA-48 | 2.600 | 2.360 | 2.840 | < 0.001*** |
| blaIMP | 0.840 | 0.560 | 1.120 | < 0.001*** |

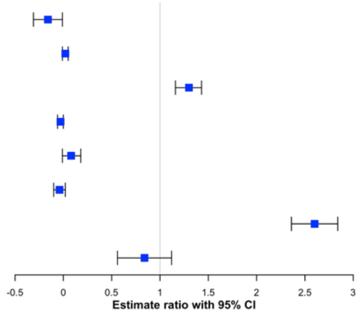

**Notes**
Results for blaVIM, blaOXA-48, and blaIMP account for only 1 collection method (passive) and 2 concentration methods (NTP and bead-beating)
-- = not applicable; all replicates had detects
OR = odds ratio (95% Confidence Interval)

**3C. Linear models predicting variance within triplicates excluding non-detects**

| Target | Est | 95% CI lower | 95% CI upper | p-value |
|---|---|---|---|---|
| C. auris | 0.140 | 0.050 | 0.220 | 0.002** |
| blaKPC | -0.030 | -0.140 | 0.080 | 0.606 |
| blaNDM | -0.170 | -0.420 | 0.070 | 0.167 |
| C.communis | 0.020 | 0.000 | 0.030 | 0.035* |
| blaVIM | 0.140 | -0.190 | 0.470 | 0.393 |
| blaOXA-48 | -0.430 | -0.740 | -0.110 | 0.019* |
| blaIMP | -0.280 | -0.450 | -0.110 | 0.002** |

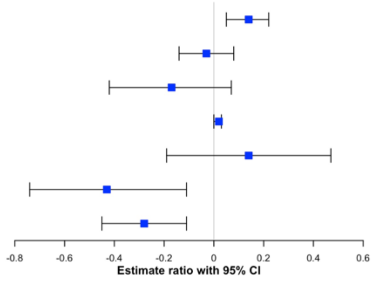

| Target | Est | 95% CI lower | 95% CI upper |
|---|---|---|---|
| C.lusitaniae | -219.530 | -353.060 | 86.010 |

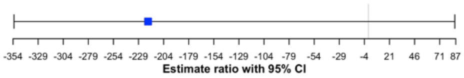

**Notes**
Results for blaVIM, blaOXA-48, and blaIMP account for only 1 collection method (passive) and 2 concentration methods (NTP and bead-beating)
Est = slope estimate (95% Confidence Interval)

Table S4. Comparison of grab and passive sampling versus composite (reference) for (A) variance with *C. lusitaniae* variance shown separately and (B) number of detections (0-3) within triplicates.

**4A. Linear models predicting variance within triplicates excluding non-detects**

| Target | Method | Est | 95% CI lower | 95% CI upper | p-value |
|---|---|---|---|---|---|
| C.auris | Grab | 0.070 | -0.180 | 0.320 | 0.582 |
| C.auris | Passive | 0.380 | 0.040 | 0.710 | 0.027* |
| blaKPC | Grab | -- | -- | -- | -- |
| blaKPC | Passive | -- | -- | -- | -- |
| blaNDM | Grab | -0.230 | -0.470 | 0.010 | 0.064 |
| blaNDM | Passive | 0.050 | -0.280 | 0.370 | 0.784 |
| C.communis | Grab | -0.030 | -0.090 | 0.030 | 0.295 |
| C.communis | Passive | -0.090 | -0.170 | -0.020 | 0.013* |
| C.lusitaniae recovery | Grab | 0.130 | 0.000 | 0.260 | 0.044* |
| C.lusitaniae recovery | Passive | -0.080 | -0.250 | 0.080 | 0.317 |
| blaVIM | Grab | 0.010 | -0.040 | 0.060 | 0.663 |
| blaVIM | Passive | -0.030 | -0.080 | 0.030 | 0.411 |
| blaOXA-48 | Grab | -0.290 | -0.490 | -0.090 | 0.004** |
| blaOXA-48 | Passive | 0.110 | -0.150 | 0.380 | 0.387 |
| blaIMP | Grab | -0.080 | -0.190 | 0.030 | 0.167 |
| blaIMP | Passive | 0.000 | -0.150 | 0.150 | 1.000 |

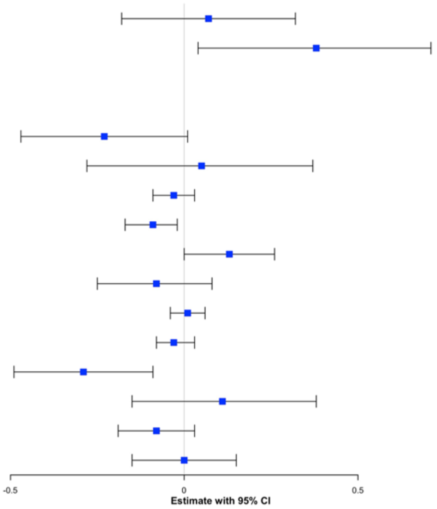

**Notes**
Est = slope estimate (95% Confidence Interval)

| Target | Method | Est | 95% CI lower | 95% CI upper | p-value |
|---|---|---|---|---|---|
| C.lusitaniae recovery | Grab | -5.990 | -14.330 | 2.340 | 0.158 |
| C.lusitaniae recovery | Passive | 1.370 | -9.730 | 12.470 | 0.808 |

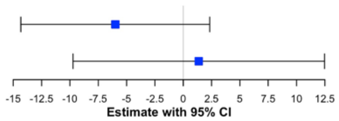

**4B. Linear models predicting number of triplicates detected**

| Target | Method | Est | 95% CI lower | 95% CI upper | p-value |
|---|---|---|---|---|---|
| C.auris | Grab | 0.070 | -0.180 | 0.320 | 0.582 |
| C.auris | Passive | 0.380 | 0.040 | 0.710 | 0.027* |
| blaKPC | Grab | -- | -- | -- | -- |
| blaKPC | Passive | -- | -- | -- | -- |
| blaNDM | Grab | -0.230 | -0.470 | 0.010 | 0.064 |
| blaNDM | Passive | 0.050 | -0.280 | 0.370 | 0.784 |
| C.communis | Grab | -0.030 | -0.090 | 0.030 | 0.295 |
| C.communis | Passive | -0.090 | -0.170 | -0.020 | 0.013* |
| C.lusitaniae recovery | Grab | 0.130 | 0.000 | 0.260 | 0.044* |
| C.lusitaniae recovery | Passive | -0.080 | -0.250 | 0.080 | 0.317 |
| blaVIM | Grab | 0.010 | -0.040 | 0.060 | 0.663 |
| blaVIM | Passive | -0.030 | -0.080 | 0.030 | 0.411 |
| blaOXA-48 | Grab | -0.290 | -0.490 | -0.090 | 0.004** |
| blaOXA-48 | Passive | 0.110 | -0.150 | 0.380 | 0.387 |
| blaIMP | Grab | -0.080 | -0.190 | 0.030 | 0.167 |
| blaIMP | Passive | 0.000 | -0.150 | 0.150 | 1.000 |

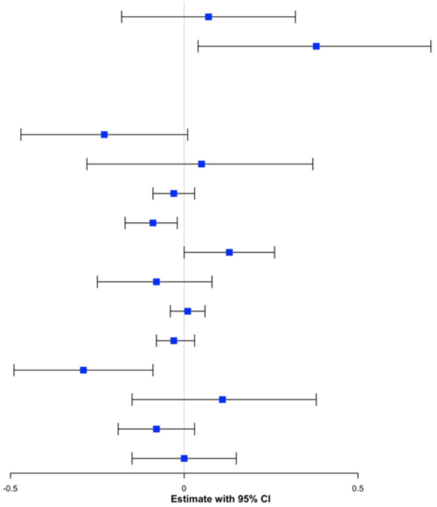

**Notes**
-- = not applicable; all available replicates had detects
Est = slope estimate (95% Confidence Interval)

Table S5. Comparison of NTP vs. ICP (Phase I) and bead-beating vs. ICP (Phase 2) for (A) variance with *C. lusitaniae* variance shown separately and (B) number of detections (0-3) within triplicates.

**5A. Linear models predicting variance within replicates excluding non-detects**

| Target | Method | Est | 95% CI lower | 95% CI upper | p-value |
|---|---|---|---|---|---|
| C.auris | Nanotrap | -0.010 | -0.070 | 0.040 | 0.620 |
| C.auris | Bead-beating | 0.150 | -0.840 | 1.140 | 0.767 |
| blaKPC | Nanotrap | -0.050 | -0.090 | -0.020 | 0.003** |
| blaKPC | Bead-beating | 0.090 | -0.410 | 0.580 | 0.728 |
| blaNDM | Nanotrap | -0.010 | -0.260 | 0.240 | 0.910 |
| blaNDM | Bead-beating | 0.010 | -0.050 | 0.070 | 0.740 |
| C.communis | Nanotrap | 0.000 | -0.050 | 0.040 | 0.902 |
| C.communis | Bead-beating | -0.050 | -0.090 | -0.020 | 0.003** |
| blaVIM | Nanotrap | -0.030 | -0.290 | 0.220 | 0.805 |
| blaVIM | Bead-beating | -0.090 | -0.140 | -0.040 | < 0.001*** |
| blaOXA-48 | Nanotrap | 0.040 | -0.000 | 0.090 | 0.052 |
| blaOXA-48 | Bead-beating | -0.060 | -0.100 | -0.020 | 0.001** |
| blaIMP | Nanotrap | -0.010 | -0.030 | 0.020 | 0.508 |
| blaIMP | Bead-beating | -0.040 | -0.070 | -0.020 | 0.002** |

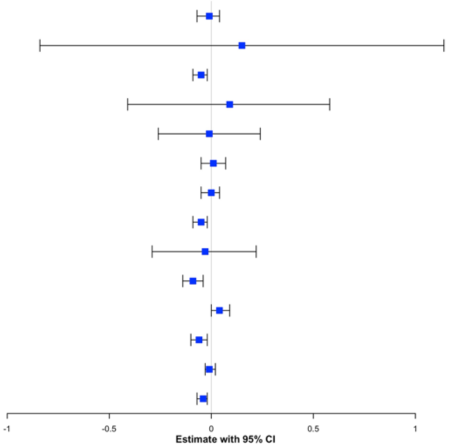

**Notes**
Est = slope estimate (95% Confidence Interval)

| Target | Method | Est | 95% CI lower | 95% CI upper | p-value |
|---|---|---|---|---|---|
| C.lusitaniae recovery | Nanotrap | 0.000 | -0.000 | 0.000 | 0.293 |
| C.lusitaniae recovery | Bead-beating | 30.170 | 10.890 | 49.440 | 0.003** |

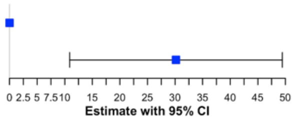

**5B. Linear models predicting number of triplicates detected**

| Target | Method | Est | 95% CI lower | 95% CI upper | p-value |
|---|---|---|---|---|---|
| C.auris | Nanotrap | -0.010 | 0.430 | 1.150 | < 0.001*** |
| C.auris | Bead-beating | 0.150 | 1.770 | 2.450 | < 0.001*** |
| blaKPC | Nanotrap | -- | -- | -- | -- |
| blaKPC | Bead-beating | -- | -- | -- | -- |
| blaNDM | Nanotrap | -0.010 | -0.600 | 0.100 | 0.155 |
| blaNDM | Bead-beating | 0.010 | 0.460 | 1.130 | < 0.001*** |
| C.communis | Nanotrap | 0.000 | -0.080 | 0.080 | 1.000 |
| C.communis | Bead-beating | -0.050 | -0.010 | 0.150 | 0.092 |
| C.lusitaniae recovery | Nanotrap | -0.030 | 0.080 | 0.380 | 0.003** |
| C.lusitaniae recovery | Bead-beating | -0.090 | -0.260 | 0.170 | 0.676 |
| blaVIM | Nanotrap | 0.040 | -0.070 | 0.070 | 1.000 |
| blaVIM | Bead-beating | -0.060 | -0.030 | 0.080 | 0.431 |
| blaOXA-48 | Nanotrap | -0.010 | -0.100 | 0.420 | 0.220 |
| blaOXA-48 | Bead-beating | -0.040 | 0.290 | 0.900 | < 0.001*** |
| blaIMP | Nanotrap | -0.010 | 0.050 | 0.370 | 0.009** |
| blaIMP | Bead-beating | 0.150 | 0.210 | 0.520 | < 0.001*** |

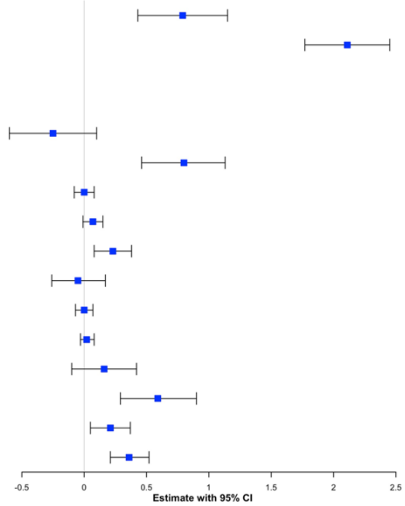

**Notes**
-- = not applicable; all replicates had detects
Est = slope estimate (95% Confidence Interval)

Table S6. Comparison of 3 categories of transport temperature (ice [reference], room, warm) for outcomes of (A) detection (yes/no), (B) number of detects (0-3), (C) variance, and (D) measured level within triplicates. PCR quantification method is dPCR.

**6A. Logistic models predicting detection/non-detection**

| Target | Transportation temp | T3P | OR | 95% CI lower | 95% CI upper | p-value |
|---|---|---|---|---|---|---|
| C.auris | Room temperature | 0.310 | 0.550 | 0.120 | 2.600 | 0.448 |
| C.auris | Warm | | 2.010 | 0.370 | 10.870 | 0.412 |
| blaKPC | Room temperature | | -- | -- | -- | -- |
| blaKPC | Warm | | -- | -- | -- | -- |
| blaNDM | Room temperature | 2.360 | 1.700 | 0.520 | 5.600 | 0.378 |
| blaNDM | Warm | | 0.550 | 0.160 | 1.950 | 0.354 |
| C.communis | Room temperature | 2.360 | 1.700 | 0.520 | 5.600 | 0.378 |
| C.communis | Warm | | 0.550 | 0.160 | 1.950 | 0.354 |
| C.lusitaniae recovery | Room temperature | 0.700 | 0.090 | 0.000 | 3.470 | 0.192 |
| C.lusitaniae recovery | Warm | | 1.000 | 0.020 | 52.970 | 1.000 |
| blaVIM | Room temperature | | -- | -- | -- | -- |
| blaVIM | Warm | | -- | -- | -- | -- |
| blaOXA-48 | Room temperature | 0.680 | 1.440 | 0.260 | 8.040 | 0.671 |
| blaOXA-48 | Warm | | 5.100 | 0.470 | 54.810 | 0.176 |
| blaIMP | Room temperature | | -- | -- | -- | -- |
| blaIMP | Warm | | -- | -- | -- | -- |

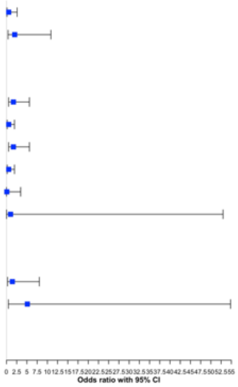

**Notes**
-- = not applicate; all replicates had detects
T3P = type 3 effects in SAS were used to generate the test with 2df

**6B. Linear models predicting number of triplicates detected**

| Target | Transportation temp | T3P | Est | 95% CI lower | 95% CI upper | p-value |
|---|---|---|---|---|---|---|
| C.auris | Room temperature | 1.56 | -0.200 | -0.670 | 0.270 | 0.385 |
| C.auris | Warm | | 0.200 | -0.270 | 0.670 | 0.385 |
| blaKPC | Room temperature | | -- | -- | -- | -- |
| blaKPC | Warm | | -- | -- | -- | -- |
| blaNDM | Room temperature | 0.77 | 0.300 | -0.280 | 0.880 | 0.292 |
| blaNDM | Warm | | -0.300 | -0.880 | 0.280 | 0.292 |
| C.communis | Room temperature | 0.134 | 0.200 | -0.030 | 0.430 | 0.083 |
| C.communis | Warm | | 0.200 | -0.030 | 0.430 | 0.083 |
| C.lusitaniae recovery | Room temperature | 0.387 | -0.200 | -0.540 | 0.140 | 0.236 |
| C.lusitaniae recovery | Warm | | -0.000 | -0.340 | 0.340 | 1.000 |
| blaVIM | Room temperature | | -- | -- | -- | -- |
| blaVIM | Warm | | -- | -- | -- | -- |
| blaOXA-48 | Room temperature | 2.66 | 0.100 | -0.450 | 0.650 | 0.708 |
| blaOXA-48 | Warm | | 0.300 | -0.250 | 0.850 | 0.268 |
| blaIMP | Room temperature | | -- | -- | -- | -- |
| blaIMP | Warm | | -- | -- | -- | -- |

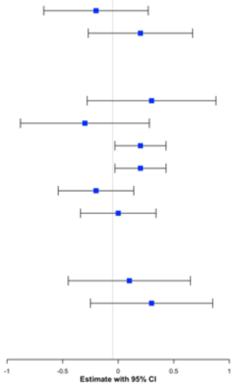

**Notes**
-- = not applicate; all replicates had detects
Est = slope estimate (95% Confidence Interval)
T3P = type 3 effects in SAS were used to generate the test with 2df

**6C. Linear models predicting variance within triplicates excluding non-detects**

| Target | Transportation temp | T3P | Est | 95% CI lower | 95% CI upper | p-value |
|---|---|---|---|---|---|---|
| C.auris | Room temperature | 0.914 | -0.060 | -0.360 | 0.230 | 0.674 |
| C.auris | Warm | | -0.040 | -0.320 | 0.240 | 0.800 |
| blaKPC | Room temperature | < 0.001*** | 0.190 | 0.070 | 0.310 | 0.003** |
| blaKPC | Warm | | 0.260 | 0.140 | 0.380 | < 0.001*** |
| blaNDM | Room temperature | 0.480 | 0.060 | -0.130 | 0.240 | 0.542 |
| blaNDM | Warm | | 0.200 | -0.000 | 0.410 | 0.055 |
| C.communis | Room temperature | 1.220 | 0.140 | -0.020 | 0.300 | 0.080 |
| C.communis | Warm | | 0.180 | 0.020 | 0.340 | 0.024* |
| C.lusitaniae recovery | Room temperature | 0.867 | -0.430 | -2.210 | 1.350 | 0.633 |
| C.lusitaniae recovery | Warm | | -0.400 | -2.180 | 1.380 | 0.656 |
| blaVIM | Room temperature | < 0.001*** | 0.210 | 0.100 | 0.310 | < 0.001*** |
| blaVIM | Warm | | 0.160 | 0.050 | 0.260 | 0.004** |
| blaOXA-48 | Room temperature | 0.077 | 0.090 | -0.060 | 0.240 | 0.240 |
| blaOXA-48 | Warm | | 0.170 | 0.020 | 0.310 | 0.024* |
| blaIMP | Room temperature | 0.092 | 0.150 | 0.000 | 0.300 | 0.049* |
| blaIMP | Warm | | 0.140 | -0.010 | 0.290 | 0.070 |

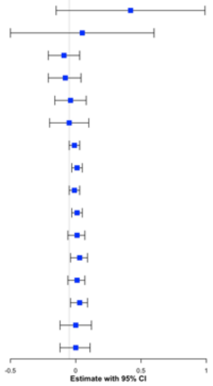

**Notes**
Est = slope estimate (95% Confidence Interval)
T3P = type 3 effects in SAS were used to generate the test with 2df

**6D. Linear models predicting level excluding non-detects**

| Target | Transportation temp | T3P | Est | 95% CI lower | 95% CI upper | p-value |
|---|---|---|---|---|---|---|
| C.auris | Room temperature | 0.914 | -0.060 | -0.360 | 0.230 | 0.674 |
| C.auris | Warm | | -0.040 | -0.320 | 0.240 | 0.800 |
| blaKPC | Room temperature | < 0.001*** | 0.190 | 0.070 | 0.310 | 0.003** |
| blaKPC | Warm | | 0.260 | 0.140 | 0.380 | < 0.001*** |
| blaNDM | Room temperature | 0.480 | 0.060 | -0.130 | 0.240 | 0.542 |
| blaNDM | Warm | | 0.200 | -0.000 | 0.410 | 0.055 |
| C.communis | Room temperature | 1.220 | 0.140 | -0.020 | 0.300 | 0.080 |
| C.communis | Warm | | 0.180 | 0.020 | 0.340 | 0.024* |
| C.lusitaniae recovery | Room temperature | 0.867 | -0.430 | -2.210 | 1.350 | 0.633 |
| C.lusitaniae recovery | Warm | | -0.400 | -2.180 | 1.380 | 0.656 |
| blaVIM | Room temperature | < 0.001*** | 0.210 | 0.100 | 0.310 | < 0.001*** |
| blaVIM | Warm | | 0.160 | 0.050 | 0.260 | 0.004** |
| blaOXA-48 | Room temperature | 0.077 | 0.090 | -0.060 | 0.240 | 0.240 |
| blaOXA-48 | Warm | | 0.170 | 0.020 | 0.310 | 0.024* |
| blaIMP | Room temperature | 0.092 | 0.150 | 0.000 | 0.300 | 0.049* |
| blaIMP | Warm | | 0.140 | -0.010 | 0.290 | 0.070 |

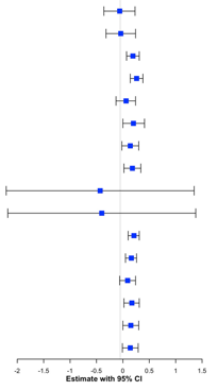

**Notes**
Linear model predicting level uses a log10 transformed quantitive outcome
Est = slope estimate (95% Confidence Interval)
T3P = type 3 effects in SAS were used to generate the test with 2df

Table S7. Comparison of 7 categories of storage for outcomes of (A) detection (yes/no), (B) number of detects (0-3), (C) variance within triplicates, and (D) measured level within triplicates.  Reference for all comparisons is immediate processing (no storage). PCR quantification method is dPCR.

### 7A. Logistic models predicting detection/non-detection

| Target | Sample storage | T3P | OR | 95% CI lower | 95% CI upper | p-value |
|---|---|---|---|---|---|---|
| C. auris | Raw 4°C - 24hr | 0.361 | 1.280 | 0.190 | 8.500 | 0.794 |
| C. auris | Raw -20°C - 7d | | 8.740 | 0.980 | 78.010 | 0.052 |
| C. auris | Raw -20°C - 1mon | | 4.930 | 0.610 | 40.140 | 0.135 |
| C. auris | Conc 4°C - 24hr | | 1.160 | 0.170 | 7.800 | 0.880 |
| C. auris | Conc -20°C - 7d | | 1.640 | 0.240 | 11.060 | 0.611 |
| C. auris | Conc -20°C - 1mon | | 1.070 | 0.160 | 7.240 | 0.943 |
| blaKPC | Raw 4°C - 24hr | | -- | -- | -- | -- |
| blaKPC | Raw -20°C - 7d | | -- | -- | -- | -- |
| blaKPC | Raw -20°C - 1mon | | -- | -- | -- | -- |
| blaKPC | Conc 4°C - 24hr | | -- | -- | -- | -- |
| blaKPC | Conc -20°C - 7d | | -- | -- | -- | -- |
| blaKPC | Conc -20°C - 1mon | | -- | -- | -- | -- |
| blaNDM | Raw 4°C - 24hr | 0.016* | 1.360 | 0.400 | 4.600 | 0.616 |
| blaNDM | Raw -20°C - 7d | | 4.150 | 1.160 | 14.830 | 0.029* |
| blaNDM | Raw -20°C - 1mon | | 14.790 | 3.080 | 70.930 | < 0.001*** |
| blaNDM | Conc 4°C - 24hr | | 1.840 | 0.550 | 6.230 | 0.322 |
| blaNDM | Conc -20°C - 7d | | 1.360 | 0.400 | 4.580 | 0.621 |
| blaNDM | Conc -20°C - 1mon | | 2.920 | 0.850 | 10.060 | 0.089 |
| C.communis | Raw 4°C - 24hr | | -- | -- | -- | -- |
| C.communis | Raw -20°C - 7d | | -- | -- | -- | -- |
| C.communis | Raw -20°C - 1mon | | -- | -- | -- | -- |
| C.communis | Conc 4°C - 24hr | | -- | -- | -- | -- |
| C.communis | Conc -20°C - 7d | | -- | -- | -- | -- |
| C.communis | Conc -20°C - 1mon | | -- | -- | -- | -- |
| C. lusitaniae recovery | Raw 4°C - 24hr | 0.713 | 2.690 | 0.120 | 61.000 | 0.532 |
| C. lusitaniae recovery | Raw -20°C - 7d | | 1.900 | 0.100 | 36.880 | 0.669 |
| C. lusitaniae recovery | Raw -20°C - 1mon | | 1.900 | 0.100 | 36.880 | 0.669 |
| C. lusitaniae recovery | Conc 4°C - 24hr | | 1.200 | 0.070 | 19.970 | 0.896 |
| C. lusitaniae recovery | Conc -20°C - 7d | | 2.490 | 0.110 | 55.170 | 0.561 |
| C. lusitaniae recovery | Conc -20°C - 1mon | | 0.320 | 0.030 | 3.840 | 0.367 |
| blaVIM | Raw 4°C - 24hr | | -- | -- | -- | -- |
| blaVIM | Raw -20°C - 7d | | -- | -- | -- | -- |
| blaVIM | Raw -20°C - 1mon | | -- | -- | -- | -- |
| blaVIM | Conc 4°C - 24hr | | -- | -- | -- | -- |
| blaVIM | Conc -20°C - 7d | | -- | -- | -- | -- |
| blaVIM | Conc -20°C - 1mon | | -- | -- | -- | -- |
| blaOXA-48 | Raw 4°C - 24hr | 0.742 | 1.730 | 0.370 | 8.090 | 0.484 |
| blaOXA-48 | Raw -20°C - 7d | | 5.840 | 0.580 | 59.230 | 0.134 |
| blaOXA-48 | Raw -20°C - 1mon | | 2.390 | 0.460 | 12.380 | 0.295 |
| blaOXA-48 | Conc 4°C - 24hr | | 1.240 | 0.290 | 5.400 | 0.769 |
| blaOXA-48 | Conc -20°C - 7d | | 2.920 | 0.450 | 18.790 | 0.257 |
| blaOXA-48 | Conc -20°C - 1mon | | 1.700 | 0.370 | 7.910 | 0.496 |
| blaIMP | Raw 4°C - 24hr | | -- | -- | -- | -- |
| blaIMP | Raw -20°C - 7d | | -- | -- | -- | -- |
| blaIMP | Raw -20°C - 1mon | | -- | -- | -- | -- |
| blaIMP | Conc 4°C - 24hr | | -- | -- | -- | -- |
| blaIMP | Conc -20°C - 7d | | -- | -- | -- | -- |
| blaIMP | Conc -20°C - 1mon | | -- | -- | -- | -- |

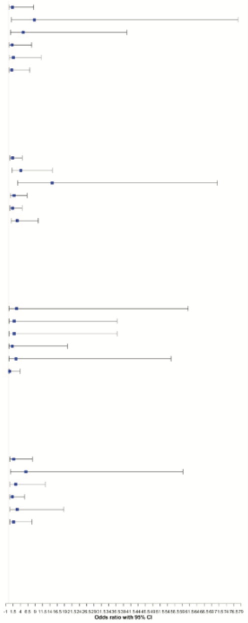

**Notes**
-- = not applicable due to lack of non-detect data
T3P = type 3 effects in SAS were used to generate the test with 2df

### 7B. Linear models predicting number of replicates detected

| Target | Sample storage | T3P | Est | 95% CI lower | 95% CI upper | p-value |
|---|---|---|---|---|---|---|
| C. auris | Raw 4°C - 24hr | 0.347 | 0.100 | -0.760 | 0.960 | 0.817 |
| C. auris | Raw -20°C - 7d | | 0.800 | -0.060 | 1.660 | 0.068 |
| C. auris | Raw -20°C - 1mon | | 0.600 | -0.260 | 1.460 | 0.169 |
| C. auris | Conc 4°C - 24hr | | 0.000 | -0.860 | 0.860 | 1.000 |
| C. auris | Conc -20°C - 7d | | 0.200 | -0.660 | 1.060 | 0.644 |
| C. auris | Conc -20°C - 1mon | | 0.000 | -0.860 | 0.860 | 1.000 |
| blaKPC | Raw 4°C - 24hr | | -- | -- | -- | -- |
| blaKPC | Raw -20°C - 7d | | -- | -- | -- | -- |
| blaKPC | Raw -20°C - 1mon | | -- | -- | -- | -- |
| blaKPC | Conc 4°C - 24hr | | -- | -- | -- | -- |
| blaKPC | Conc -20°C - 7d | | -- | -- | -- | -- |
| blaKPC | Conc -20°C - 1mon | | -- | -- | -- | -- |
| blaNDM | Raw 4°C - 24hr | 0.008** | 0.200 | -0.560 | 0.960 | 0.601 |
| blaNDM | Raw -20°C - 7d | | 0.900 | 0.140 | 1.660 | 0.022* |
| blaNDM | Raw -20°C - 1mon | | 1.400 | 0.640 | 2.160 | < 0.001*** |
| blaNDM | Conc 4°C - 24hr | | 0.400 | -0.360 | 1.160 | 0.298 |
| blaNDM | Conc -20°C - 7d | | 0.200 | -0.560 | 0.960 | 0.601 |
| blaNDM | Conc -20°C - 1mon | | 0.700 | -0.060 | 1.460 | 0.071 |
| C.communis | Raw 4°C - 24hr | | -- | -- | -- | -- |
| C.communis | Raw -20°C - 7d | | -- | -- | -- | -- |
| C.communis | Raw -20°C - 1mon | | -- | -- | -- | -- |
| C.communis | Conc 4°C - 24hr | | -- | -- | -- | -- |
| C.communis | Conc -20°C - 7d | | -- | -- | -- | -- |
| C.communis | Conc -20°C - 1mon | | -- | -- | -- | -- |
| C. lusitaniae recovery | Raw 4°C - 24hr | 0.523 | 0.200 | -0.430 | 0.830 | 0.524 |
| C. lusitaniae recovery | Raw -20°C - 7d | | 0.100 | -0.530 | 0.730 | 0.750 |
| C. lusitaniae recovery | Raw -20°C - 1mon | | 0.100 | -0.530 | 0.730 | 0.750 |
| C. lusitaniae recovery | Conc 4°C - 24hr | | 0.000 | -0.630 | 0.630 | 1.000 |
| C. lusitaniae recovery | Conc -20°C - 7d | | 0.200 | -0.430 | 0.830 | 0.524 |
| C. lusitaniae recovery | Conc -20°C - 1mon | | -0.400 | -1.030 | 0.230 | 0.206 |
| blaVIM | Raw 4°C - 24hr | | -- | -- | -- | -- |
| blaVIM | Raw -20°C - 7d | | -- | -- | -- | -- |
| blaVIM | Raw -20°C - 1mon | | -- | -- | -- | -- |
| blaVIM | Conc 4°C - 24hr | | -- | -- | -- | -- |
| blaVIM | Conc -20°C - 7d | | -- | -- | -- | -- |
| blaVIM | Conc -20°C - 1mon | | -- | -- | -- | -- |
| blaOXA-48 | Raw 4°C - 24hr | 0.706 | 0.200 | -0.300 | 0.700 | 0.430 |
| blaOXA-48 | Raw -20°C - 7d | | 0.430 | -0.090 | 0.950 | 0.100 |
| blaOXA-48 | Raw -20°C - 1mon | | 0.300 | -0.200 | 0.800 | 0.238 |
| blaOXA-48 | Conc 4°C - 24hr | | 0.100 | -0.400 | 0.600 | 0.692 |
| blaOXA-48 | Conc -20°C - 7d | | 0.320 | -0.200 | 0.840 | 0.219 |
| blaOXA-48 | Conc -20°C - 1mon | | 0.200 | -0.300 | 0.700 | 0.430 |
| blaIMP | Raw 4°C - 24hr | | -- | -- | -- | -- |
| blaIMP | Raw -20°C - 7d | | -- | -- | -- | -- |
| blaIMP | Raw -20°C - 1mon | | -- | -- | -- | -- |
| blaIMP | Conc 4°C - 24hr | | -- | -- | -- | -- |
| blaIMP | Conc -20°C - 7d | | -- | -- | -- | -- |
| blaIMP | Conc -20°C - 1mon | | -- | -- | -- | -- |

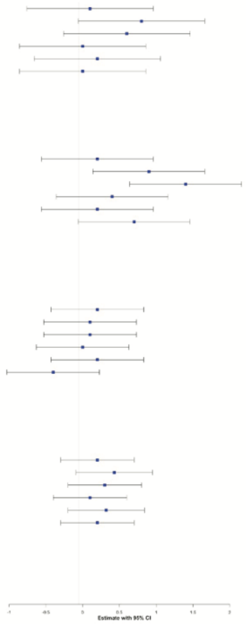

**Notes**
-- = not applicable due to lack of non-detect data
Est = slope estimate (95% Confidence Interval)
T3P = type 3 effects in SAS were used to generate the test with 2df

### 7C. Linear models predicting variance within triplicates excluding non-detects

| Target | Sample storage | T3P | Est | 95% CI lower | 95% CI upper | p-value |
|---|---|---|---|---|---|---|
| C. auris | Raw 4°C - 24hr | 0.103 | -0.530 | -0.960 | -0.100 | 0.017* |
| C. auris | Raw -20°C - 7d | | -0.510 | -0.910 | -0.110 | 0.013* |
| C. auris | Raw -20°C - 1mon | | -0.440 | -0.840 | -0.040 | 0.034* |
| C. auris | Conc 4°C - 24hr | | -0.450 | -0.890 | 0.000 | 0.051 |
| C. auris | Conc -20°C - 7d | | -0.630 | -1.070 | -0.190 | 0.006** |
| C. auris | Conc -20°C - 1mon | | -0.580 | -1.010 | -0.150 | 0.010* |
| blaKPC | Raw 4°C - 24hr | 0.532 | 0.020 | -0.200 | 0.240 | 0.867 |
| blaKPC | Raw -20°C - 7d | | 0.140 | -0.080 | 0.370 | 0.204 |
| blaKPC | Raw -20°C - 1mon | | 0.070 | -0.160 | 0.290 | 0.547 |
| blaKPC | Conc 4°C - 24hr | | -0.020 | -0.240 | 0.210 | 0.875 |
| blaKPC | Conc -20°C - 7d | | 0.160 | -0.070 | 0.380 | 0.161 |
| blaKPC | Conc -20°C - 1mon | | 0.130 | -0.090 | 0.350 | 0.249 |
| blaNDM | Raw 4°C - 24hr | 0.379 | 0.040 | -0.060 | 0.140 | 0.426 |
| blaNDM | Raw -20°C - 7d | | 0.010 | -0.080 | 0.100 | 0.846 |
| blaNDM | Raw -20°C - 1mon | | 0.020 | -0.070 | 0.100 | 0.709 |
| blaNDM | Conc 4°C - 24hr | | 0.010 | -0.090 | 0.100 | 0.914 |
| blaNDM | Conc -20°C - 7d | | 0.000 | -0.100 | 0.100 | 0.977 |
| blaNDM | Conc -20°C - 1mon | | 0.070 | -0.020 | 0.160 | 0.115 |
| C.communis | Raw 4°C - 24hr | 0.668 | -0.020 | -0.140 | 0.110 | 0.800 |
| C.communis | Raw -20°C - 7d | | 0.030 | -0.090 | 0.150 | 0.628 |
| C.communis | Raw -20°C - 1mon | | 0.040 | -0.080 | 0.160 | 0.535 |
| C.communis | Conc 4°C - 24hr | | -0.060 | -0.180 | 0.060 | 0.341 |
| C.communis | Conc -20°C - 7d | | -0.020 | -0.140 | 0.110 | 0.802 |
| C.communis | Conc -20°C - 1mon | | 0.040 | -0.090 | 0.160 | 0.548 |
| C. lusitaniae recovery | Raw 4°C - 24hr | 0.668 | -0.020 | -0.140 | 0.110 | 0.800 |
| C. lusitaniae recovery | Raw -20°C - 7d | | 0.030 | -0.090 | 0.150 | 0.628 |
| C. lusitaniae recovery | Raw -20°C - 1mon | | 0.040 | -0.080 | 0.160 | 0.535 |
| C. lusitaniae recovery | Conc 4°C - 24hr | | -0.060 | -0.180 | 0.060 | 0.341 |
| C. lusitaniae recovery | Conc -20°C - 7d | | -0.020 | -0.140 | 0.110 | 0.802 |
| C. lusitaniae recovery | Conc -20°C - 1mon | | 0.040 | -0.090 | 0.160 | 0.548 |
| blaVIM | Raw 4°C - 24hr | 0.288 | 1.030 | 0.070 | 2.000 | 0.036* |
| blaVIM | Raw -20°C - 7d | | -0.030 | -1.020 | 0.970 | 0.960 |
| blaVIM | Raw -20°C - 1mon | | -0.010 | -0.970 | 0.960 | 0.985 |
| blaVIM | Conc 4°C - 24hr | | 0.060 | -0.900 | 1.030 | 0.897 |
| blaVIM | Conc -20°C - 7d | | 0.040 | -0.950 | 1.040 | 0.929 |
| blaVIM | Conc -20°C - 1mon | | 0.050 | -0.910 | 1.020 | 0.916 |
| blaOXA-48 | Raw 4°C - 24hr | 0.288 | 1.030 | 0.070 | 2.000 | 0.036* |
| blaOXA-48 | Raw -20°C - 7d | | -0.030 | -1.020 | 0.970 | 0.960 |
| blaOXA-48 | Raw -20°C - 1mon | | -0.010 | -0.970 | 0.960 | 0.985 |
| blaOXA-48 | Conc 4°C - 24hr | | 0.060 | -0.900 | 1.030 | 0.897 |
| blaOXA-48 | Conc -20°C - 7d | | 0.040 | -0.950 | 1.040 | 0.929 |
| blaOXA-48 | Conc -20°C - 1mon | | 0.050 | -0.910 | 1.020 | 0.916 |
| blaIMP | Raw 4°C - 24hr | 0.454 | 0.170 | -0.090 | 0.440 | 0.197 |
| blaIMP | Raw -20°C - 7d | | -0.050 | -0.310 | 0.210 | 0.707 |
| blaIMP | Raw -20°C - 1mon | | -0.030 | -0.290 | 0.230 | 0.824 |
| blaIMP | Conc 4°C - 24hr | | 0.170 | -0.090 | 0.430 | 0.198 |
| blaIMP | Conc -20°C - 7d | | 0.000 | -0.260 | 0.260 | 0.994 |
| blaIMP | Conc -20°C - 1mon | | 0.060 | -0.200 | 0.320 | 0.624 |

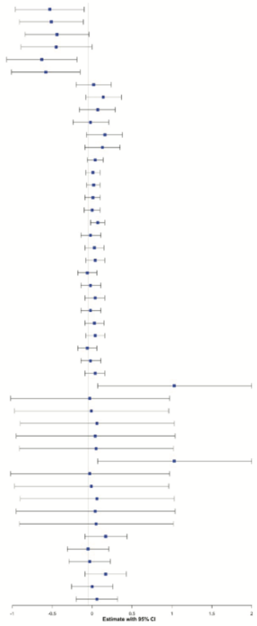

**Notes**
Linear model predicting level uses a log10 transformed quantitive outcome
Est = slope estimate (95% Confidence Interval)
T3P = type 3 effects in SAS were used to generate the test with 2df

### 7D. Linear models predicting level excluding non-detects

| Target | Sample storage | T3P | Est | 95% CI lower | 95% CI upper | p-value |
|---|---|---|---|---|---|---|
| C. auris | Raw 4°C - 24hr | < 0.001*** | -0.380 | -0.720 | -0.030 | 0.035* |
| C. auris | Raw -20°C - 7d | | -0.420 | -0.730 | -0.100 | 0.011* |
| C. auris | Raw -20°C - 1mon | | -0.530 | -0.860 | -0.210 | 0.001** |
| C. auris | Conc 4°C - 24hr | | 0.460 | 0.110 | 0.810 | 0.009** |
| C. auris | Conc -20°C - 7d | | 0.550 | 0.200 | 0.890 | 0.002** |
| C. auris | Conc -20°C - 1mon | | 0.470 | 0.120 | 0.810 | 0.009** |
| blaKPC | Raw 4°C - 24hr | 0.405 | 0.040 | -0.290 | 0.370 | 0.815 |
| blaKPC | Raw -20°C - 7d | | 0.230 | -0.100 | 0.560 | 0.165 |
| blaKPC | Raw -20°C - 1mon | | 0.020 | -0.310 | 0.350 | 0.896 |
| blaKPC | Conc 4°C - 24hr | | 0.320 | -0.010 | 0.650 | 0.055 |
| blaKPC | Conc -20°C - 7d | | 0.170 | -0.160 | 0.500 | 0.310 |
| blaKPC | Conc -20°C - 1mon | | 0.160 | -0.170 | 0.480 | 0.354 |
| blaNDM | Raw 4°C - 24hr | < 0.001*** | -0.250 | -0.590 | 0.080 | 0.136 |
| blaNDM | Raw -20°C - 7d | | 0.260 | -0.040 | 0.560 | 0.090 |
| blaNDM | Raw -20°C - 1mon | | 0.390 | 0.100 | 0.680 | 0.009** |
| blaNDM | Conc 4°C - 24hr | | 0.410 | 0.100 | 0.730 | 0.011* |
| blaNDM | Conc -20°C - 7d | | 0.420 | 0.080 | 0.750 | 0.015* |
| blaNDM | Conc -20°C - 1mon | | 0.390 | 0.090 | 0.700 | 0.013* |
| C.communis | Raw 4°C - 24hr | 0.007** | 0.050 | -0.350 | 0.450 | 0.795 |
| C.communis | Raw -20°C - 7d | | 0.550 | 0.160 | 0.950 | 0.007** |
| C.communis | Raw -20°C - 1mon | | 0.080 | -0.310 | 0.480 | 0.676 |
| C.communis | Conc 4°C - 24hr | | 0.030 | -0.370 | 0.430 | 0.894 |
| C.communis | Conc -20°C - 7d | | 0.250 | -0.150 | 0.640 | 0.221 |
| C.communis | Conc -20°C - 1mon | | -0.260 | -0.660 | 0.130 | 0.194 |
| C. lusitaniae recovery | Raw 4°C - 24hr | 0.195 | 1.740 | -0.730 | 4.210 | 0.165 |
| C. lusitaniae recovery | Raw -20°C - 7d | | 2.320 | -0.150 | 4.790 | 0.065 |
| C. lusitaniae recovery | Raw -20°C - 1mon | | 2.270 | -0.200 | 4.740 | 0.071 |
| C. lusitaniae recovery | Conc 4°C - 24hr | | 0.190 | -2.280 | 2.660 | 0.877 |
| C. lusitaniae recovery | Conc -20°C - 7d | | -0.040 | -2.510 | 2.430 | 0.977 |
| C. lusitaniae recovery | Conc -20°C - 1mon | | 0.530 | -1.940 | 3.000 | 0.674 |
| blaVIM | Raw 4°C - 24hr | < 0.001*** | -0.420 | -0.730 | -0.110 | 0.008** |
| blaVIM | Raw -20°C - 7d | | 0.360 | 0.050 | 0.680 | 0.025* |
| blaVIM | Raw -20°C - 1mon | | 0.360 | 0.060 | 0.670 | 0.021* |
| blaVIM | Conc 4°C - 24hr | | -0.030 | -0.340 | 0.270 | 0.823 |
| blaVIM | Conc -20°C - 7d | | -0.090 | -0.410 | 0.220 | 0.557 |
| blaVIM | Conc -20°C - 1mon | | 0.010 | -0.300 | 0.320 | 0.951 |
| blaOXA-48 | Raw 4°C - 24hr | < 0.001*** | 2.240 | 1.880 | 2.600 | < 0.001*** |
| blaOXA-48 | Raw -20°C - 7d | | 2.380 | 2.020 | 2.740 | < 0.001*** |
| blaOXA-48 | Raw -20°C - 1mon | | 1.850 | 1.500 | 2.210 | < 0.001*** |
| blaOXA-48 | Conc 4°C - 24hr | | -0.410 | -0.780 | -0.050 | 0.027* |
| blaOXA-48 | Conc -20°C - 7d | | -0.790 | -1.150 | -0.420 | < 0.001*** |
| blaOXA-48 | Conc -20°C - 1mon | | -0.390 | -0.750 | -0.030 | 0.035* |
| blaIMP | Raw 4°C - 24hr | < 0.001*** | 2.150 | 1.820 | 2.480 | < 0.001*** |
| blaIMP | Raw -20°C - 7d | | 2.590 | 2.270 | 2.920 | < 0.001*** |
| blaIMP | Raw -20°C - 1mon | | 2.460 | 2.140 | 2.780 | < 0.001*** |
| blaIMP | Conc 4°C - 24hr | | 2.180 | 1.850 | 2.500 | < 0.001*** |
| blaIMP | Conc -20°C - 7d | | 1.850 | 1.520 | 2.170 | < 0.001*** |
| blaIMP | Conc -20°C - 1mon | | 2.220 | 1.890 | 2.550 | < 0.001*** |

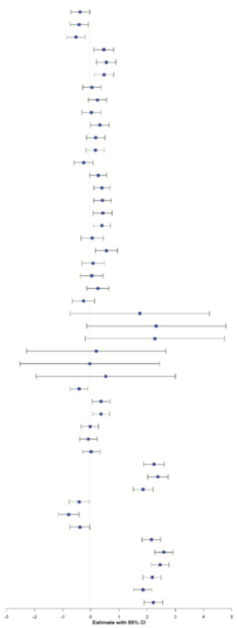

**Notes**
Linear model predicting level uses a log10 transformed quantitive outcome
Est = slope estimate (95% Confidence Interval)
T3P = type 3 effects in SAS were used to generate the test with 2df

**Supplemental Methods**

**Visual tracer dye test**

To visually assess that the sewer represented all facility wastewater, we used 10g of yellow-green powder leak tracer dye (Bright Dyes® FLT Yellow-Green Powder) dissolved in 500mL of deionized water in a 1L Nalgene bottle. Four bottles were prepared for the test. On-site, we selected the floor nearest and farthest from the manhole, as identified by the facilities management team. The dye was then flushed in two bathrooms per floor, one in the north wing and one in the south wing. An outside team monitored the manhole using an LED or UV

flashlight, recording the times of (1) the toilet flush, (2) the first visual detection of the

tracer, (3) an increase or decrease in flow, and (4) the end of visual detection of the tracer, as

well as recording the number of flushes or if the sink or showers were turned on during the evaluation.

**Sample collection**

Time-weighted composite samples were collected over a 24h using an AS950 portable autosampler (Hach) programmed to collected 400mL every 2h. 1 L of the composite sample was collected for processing. Before aliquoting 1L of sample into a sterile bottle, the collection carboy was thoroughly shaken to homogenize the sample and ensure solids were not settled at the bottom. To maintain sample integrity and prevent cross-contamination, the Tygon tubing used for the autosampler was replaced every 1-2 weeks, as cleaning alone was insufficient to remove accumulated wastewater residuals and biofilm. Additionally, the sampler was thoroughly cleaned with 70% isopropyl alcohol during each visit. The carboy was cleaned weekly by soaking with Bleach-Rite hospital-grade bleach disinfectant for 24h and rinsed with deionized water.

Grab samples were collected using either a swing sampler (Nisco) attached to a 1L sterile polyethylene bottle, secured with a snapper ring, or with the AS950 autosampler configured to manual mode.

Passive samples were obtained using three 4.5 x 4.1 yd sterile cotton gauze bandages (Dukal), which were placed in a stainless-steel cage (Fig. S1) and suspended from a stainless-steel cable attached to a custom-designed brace at the top of the manhole (Fig. S1). The gauze was placed 24 h prior to the scheduled collection. At retrieval, the samples were placed in sterile plastic bags. In the laboratory, ~150 mL wastewater was extracted from the gauze by squeezing them inside the plastic bag and decanting into sterile 50mL conical tubes.

Temperature and flow measurements were logged hourly for 24 h preceding sample collection using a FL-16 Global Logger (400752-25, YSI). Turbidity and pH measurements were taken during sample collection using a Multiparameter Meter (HI9829, Hanna Instruments) with the following attachments; autonomously logging probe (HI7629829, Hanna Instruments), pH probe (HI7629829-1, Hanna Instruments), turbidity probe (HI7629829-4, Hanna Instruments). Turbidity and pH were measured every 30 s for 10 min during sample retrieval.

### Concentration

For the magnetic bead-based extraction, 75 µL Microbiome B Nanotrap particles + Enhancement Reagent 3 (Ceres) were added to 10mL wastewater with 50µL of added *C. lusitaniae*. Automated concentration in 24-deep-well plates was done as specified by the manufacturer using the Kingfisher Apex (ThermoFisher). Samples were eluted into 500 µL of MagMAX Microbiome Lysis Solution (ThermoFisher).

Filtration-based concentration used the InnovaPrep Concentrating Pipette Select (ICP). For the ICP method, 50µL of *C.lusitaniae* were added to each 50mL wastewater aliquot and then vortexed for 30 seconds to homogenize. Next, samples were centrifuged at 4°C for 10 minutes at 3000 x *g* to pellet the solids. The supernatant was transferred to a 50mL tube and filtered through a 0.45-µm pore-size tip to capture particles. The retained particles in the filter tip were eluted in 500-1000µL using the manufacturer's wet foam elution canister.

For the centrifugation and bead-beating approach, 50µL of *C. lusitaniae* were added to each 50mL of wastewater aliquot which was then centrifuged at 4°C for 10 minutes at 3000 x *g* to pellet the solids. After centrifugation, the supernatant was discarded, and a 200-300mg pellet was transferred to a 15mL conical centrifuge tube containing 0.5g of 0.5mm zirconia/silica beads (BioSpec) and bead-beat using the FastPrep-24 (MP Biomedicals) at 4.0 m/s for 60 s.

### DNA extraction

To extract DNA, we used the MagMAX Microbiome Lysis Solution (ThermoFisher) automated approach on the Kingfisher Apex (ThermoFisher). 92µL of Proteinase K were added to each well on a 96-deep-well plate along with 520µL of a 25:1 mixture of MagMAX Binding Solution and MagMAX DNA/RNA binding beads and 400µL of wastewater concentrate. DNA was eluted in 112µL of MagMAX elution solution.

**PCR assays**

For qPCR, CDC primers were used $bla_{KPC}$, $bla_{NDM}$, $bla_{VIM}$, and $bla_{OXA\text{-}48\text{-}like}$ and from previous studies for $bla_{IMP}$ [1], *C. auris* [2], and crAssphage [3]. Multiplexed quantitative PCR analyses for *C. auris* and carbapenamase genes were performed according to established CDC protocols (KPC/NDM/16S = ARC.TE.C.0022; OXA-48/VIM/16S = ARC.TE.C.0143) and in-development protocols (IMP/16S). Primer and probe sequences are shown in **Table S2**. All standards were performed in technical triplicate and samples were analyzed in technical duplicate. Synthetic DNA standards, synthesized as gBLOCKs (IDT) of 374 to 813 bp, were diluted across the dynamic range of each assay. Synthetic DNA sequences and sequence sources are shown in **Table S2**. Known antibiotic resistance gene-containing microorganisms were ordered from the CDC and FDA Antimicrobial Resistance Isolate Bank. All assays were performed in 384-well plates (MicroAmp™ Optical 384-Well Reaction Plate with Barcode, Applied Biosystems, cat no. #4309849) with optical adhesive covers (Applied Biosystems, cat no. #4311971), and using a QuantStudio 5 real-time PCR system (Thermo Fisher). PCR master mixes varied by assay, and included PerfeCta MultiPlex qPCR ToughMix, low ROX (Quanta Biosciences; Cat# 95149-250) or KiCqStart Probe qPCR ReadyMix, LOW ROX (Sigma cat#KCQS05). Analysis of C. auris, C. lusitaniae, for

CrAssphage, BICOID spike-in, and IMP genes were conducted using PerfeCta MultiPlex qPCR ToughMix. Analyses for VIM, OXA-48-like, KPC, and NMD carbapenemase genes were performed using the KiCqStart Probe qPCR ReadyMix. All reactions were performed in 20 microliter volumes. Nuclease free water (2-ml aliquots) were obtained from IDT (cat# 11-04-02-01). All cycling conditions were 95°C denaturation followed by 35-45 cycles of PCR with two-step cycling of 95°C for 3 seconds and 60°C for 30 seconds.

For dPCR, we used CDC-approved assays optimized for dPCR (GT-Digital AMR and DNA Pathogen Wastewater Surveillance Panel v1.0 and GT-Digital *C. auris* Wastewater Surveillance Assay Kit for the QIAGEN QIAcuity® Digital PCR System https://pmc.ncbi.nlm.nih.gov/articles/PMC10467632/ v2.0) according to manufacturer's specifications on the QIAcuity (Qiagen) digital PCR system with 2µL template or water control and 10 µL of Qiagen QIAcuity Probe PCR Kit mastermix on a QIAcuity 26k 24-well nanoplate. The results from each nanoplate were visually inspected, and the threshold was determined using the signal of the positive control, where clear separation between partitions was observed, as the default.

All dPCR was performed on the Qiagen QIAcuity platform, using assays from GT Molecular. Reaction volumes for AMR targets ($bla_{KPC}$, $bla_{NDM}$, $bla_{VIM}$, $bla_{OXA-48}$ and $bla_{IMP}$), and *C. auris*/C. *lusitaniae* were identical. dPCR reaction volumes were calculated per the GT Molecular protocols.

| Target/Kit | Reagent | Volume per reaction (µL) |
|---|---|---|

| | | |
|---|---|---|
| VIM:OXA-48, KPC:NDM, IMP, *C. auris*:*C. lusitaniae* | 4x QIAcuity Probe MasterMix (Qiagen) | 10 |
| | GT Molecular Assay Solution (GT Molecular) | 2 |
| | Molecular Water (ThermoFisher) | 18 |
| | Sample template | 10* |
| Carjivirus | 4x QIAcuity Probe MasterMix (Qiagen) | 3 |
| | GT Molecular Assay Solution (GT Molecular) | 0.6 |
| | Molecular Water (ThermoFisher) | 6.4 |
| | Sample template | 2 |

**Limit of detection (LoD)**

To determine the limit of detection for the gene targets *blaNDM, blaKPC*, *blaIMP*, *blaOXA-48*, *blaIMP*, *C.auris,* and *C.lusinatiae,* a serial dilution was performed for each biomarker [get exact volume from Chi-Yu. These targets were assayed on a QIAcuity 26k-24 well nanoplate (QIAGEN), with a reaction volume of 40 µL per well. Each plate was processed with a non-amplification negative control consisting of 10 µL of molecular-grade water, and 30 µL of master mix. For the fecal indicator (*Carjivirus*) quantification, a QIAcuity 8.5k-96 well nanoplate was used with a reaction volume of 12µL. We deemed one positive partition to be sufficient to categorize the sample as a 'detect', as all sample collections were processed in triplicate. The LoD in starting wastewater was calculated following the steps outlined in the calculations below:

1. $\left(QIAcuity\ concentration\ output\ gene\ \frac{copies}{\mu L}\right)(PCR\ reaction\ volume\ 40\mu L\ or\ 12\mu L) =$

   $Total\ gene\ copies\ in\ PCR\ reaction$

2. $\frac{Total\ gene\ copies\ in\ PCR}{Volume\ of\ total\ extracted\ nucleic\ acids\ (TNA)\ added\ to\ the\ PCR} = gene\ copies\ in\ 1\mu L\ of\ \ TNA$

3. $(Gene\ copies\ in\ 1\mu L\ of\ TNA)(Total\ extraction\ elution\ volume) = Total\ gene\ copies\ in\ volume\ extracted$

4. $\frac{Total\ gene\ copies\ in\ volume\ extracted}{Concentrate\ volume\ added\ to\ the\ extraction\ plate} = gene\ copies\ in\ 1\mu L\ of\ \ concentrate$

5. $(Gene\ copies\ in\ 1\mu L\ of\ \ concentrate)(Total\ concentrate\ volume) = Total\ gene\ copies\ in\ concentrate\ volume$

6. $\frac{Total\ gene\ copies\ in\ concentrate\ volume*1000}{Wastewater\ input\ volume} = gene\ copies/L$

The calculated LoDs were:

| Biomarker | gc/µL | LoD gc/L of WW |
|---|---|---|
| *blaNDM* | 0.053 | 1987.5 |
| *blaKPC* | 0.053 | 1987.5 |
| *blaOXA-48* | 0.054 | 2025 |
| *blaVIM* | 0.053 | 2025 |
| *blaIMP* | 0.054 | 1987.5 |

| | | |
|---|---|---|
| *C.auris* | 0.056 | 2100 |
| *C.lusitaniae* | 0.056 | 2100 |
| *Carjivirus* | 0.365 | 20531.25 |

**Process control *C. lusitaniae***

*Clavispora lusitaniae Rodrigues de Miranda* strain (ATCC 201083), was used as the process control per the GT Molecular *C. auris* assay protocol. Lyophilized cell pellets were resuspended in 1000 µL of tris-EDTA buffer (pH 8) and diluted with 9000µL of molecular-grade water to a 1:10 concentration stock. This stock was then used during the concentration step as the process control. Due to ongoing supply issues ATCC 201083 being placed on back order for 6+ months, and additional freeze-dried ampoules being unobtainable, another solution was needed for the process control. To mitigate further issues with product availability, *C. lusitaniae van Uden et do Carmo Sousa* (ATCC 34449) freeze dried pellets were resuspended per ATCC recommendations and cultured. After incubation, cultures were pelleted into 50 mL conical tubes and kept frozen at –80°C for use as the process control.

Gene copies of the spike-in were determined using heat lysis on 30µL from the resuspended stock in triplicate on a Bio-Rad C1000 Touch Thermal Cycler with the following cycling conditions: denaturation at 95°C for 10 minutes, followed by a 5-minute hold at 4°C. Next, 10µL of each heat-lysed tube were transferred to a dPCR nanoplate for quantification using the *C. lusitaniae* assay in triplicate. The following calculations were used to determine *C. lusitaniae* recovery:

$$\frac{\left(QIAcuity\ output\ \frac{gene\ copies}{\mu L}\right)(PCR\ volume)}{(Suspension\ template\ volume)(Volume\ of\ suspension\ added\ to\ each\ sample)}$$

$$= Total\ copies\ added\ to\ the\ wastewater\ sample$$

$$\frac{Total\frac{gene\ copies}{\mu L}\ detected\ in\ each\ sample}{Total\ gene\frac{copies}{\mu L}added\ to\ the\ sample*1000} = C.lusitaniae\ recovery\ percent$$

**Cultivation of CPOs and *C. auris***

A 50 mL aliquot of a wastewater composite sample was centrifuged at 4,500*xg* for 10 minutes. The supernatant was removed and the remaining 1mL eluent was homogenized using a vortex. [4]. For CPO cultivation, 100 uL was transferred to 2 mL of tryptic soy broth (Remel) with a 10 ug ertapenem disk. Broth cultures were incubated at 35°C in ambient air for 24 hours, then subcultured to mSuperCARBA (CHROMagar, Saint-Denis, France) [5]. *C. auris* was cultured according to previously published methods [6] and CDC protocol FRL-100-P03 (https://www.cdc.gov/candida-auris/hcp/laboratories/isolation-procedure.html): 100 uL of the homogenized eluent was transferred to 2 mL Sabouraud Salt Dulcitol broth with chloramphenicol and gentamicin (SSD broth) and incubated at 40°C with shaking at 250 RPM. After 7 days or earlier if broths were noted to be cloudy, 10 uL was subcultured onto CHROMagar Candida plates (Becton Dickinson; Franklin Lakes, NJ) [6]. Unique colony morphologies consistent with CPOs or *C.*

*auris* were identified to species level using MALDI-TOF mass spectroscopy (bioMérieux, Marcy-I'Étoile, France). Presumptive CPOs were tested for carbapenemase genes using the Xpert® Carba-R system (Cepheid; Sunnyvale, CA). All cultivation was done in triplicate with appropriate controls.

**Shotgun Metagenomics**

Metagenomic analysis was performed on raw wastewater, enrichment broth cultures, and selective plating cultures. Raw wastewater was prepared as follows: approximately 1 L of a grab wastewater was stored at refrigeration (2-8°C) and processed on the day of collection, usually within 2 h of receipt. Triplicate 50 mL of wastewater samples were concentrated by centrifugation for 20 min at 2,500*xg* [4]. For each triplicate, the pellet was resuspended in 1 mL of residual wastewater. For sequencing enrichment broth and selective plating culture (from above cultivation methods), we used 1 mL of cultured broth and a 10 uL loop sweep of the whole plate, each respectively added to a microcentrifuge tube with 1 mL water. All samples were agitated using a vortex, heat shocked, spun down, and then frozen until extraction.

Microbial DNA was extracted using a QIAamp PowerFecal Pro DNA Kit (Qiagen), and libraries were prepared using a Nextera XT DNA Library Preparation Kit (Illumina). Pooled libraries were sequenced on a NovaSeq X instrument, employing paired-end 2x150 base reads. Data analysis is described in the supplemental methods.

**Metagenomic sequence data analysis**

For taxonomic classification, raw reads were adapter-trimmed and quality filtered using BBDuk v38.86 and taxa assigned using MetaPhlAn v4.0.1 [7]. AMR genes were annotated on the Chan-Zuckerberg ID portal with the Antimicrobial Resistance pipeline v1.4. Briefly, reads were adapter-trimmed and quality-filtered, and human reads were removed using alignments against reference human genomes. Following duplicate removal, reads were subsampled to 2 million paired reads per sample. The Resistance Gene Identifier (RGI) tool and Comprehensive Antimicrobial Resistance Database (CARD) were used to identify resistance genes in reads and assembled contigs [8]. Identified AMR genes were quality filtered using read_coverage_breadth ≥ 95. Gene detection is reported as the relative abundance metric reads depth per million (dPM), the number of bases mapped to the reference sequence in CARD divided by the sequence length, per million reads sequenced.
